# Proactive case-finding and risk-stratification in people at risk of chronic liver disease in Greater Manchester: a cost-effectiveness analysis

**DOI:** 10.1101/2025.06.01.25328671

**Authors:** Gabriel Rogers, Stephanie Landi, Huw Purssell, Tonia Momoh, Sol Yates, Oliver Street, Karen Piper Hanley, Neil Hanley, Varinder Athwal, Katherine Payne

## Abstract

**Background:** We urgently need innovative strategies to combat a growing epidemic of chronic liver disease (CLD). ID-LIVER was a collaborative project aiming to improve detection of reversible-stage CLD in a region with high prevalence of critical risk factors.

**Objective:** To determine the cost-effectiveness of ways to identify people with significant CLD, including proactive case-finding in the community (supplementing reactive referrals from primary care) and/or risk-stratification (using FIB-4 or ID-LIVER-ML -- a novel machine-learning risk-stratification tool).

**Design:** State-transition decision-analytic model estimating lifetime healthcare costs (2023/24 GBP) and quality-adjusted life-years (QALYs) associated with six alternative strategies for case-finding and risk-stratification. We simulated cohorts of people with alcohol-related liver disease (ARLD) and metabolic dysfunction-associated steatotic liver disease (MASLD). We populated the model with data collected in ID-LIVER, supplemented by parameters from literature and routine data-sources. We estimated incremental cost-effectiveness and performed deterministic and probabilistic sensitivity analyses.

**Results:** Any case-identification strategy costing ≤3,300 GBP per person with significant CLD identified would meet English cost-effectiveness thresholds (20,000 GBP/QALY). In our decision-set, the cheapest strategy is to use FIB-4 in the reactive-only population. ID-LIVER-ML generates more population health at reasonable cost (10,490 GBP/QALY gained). Introducing proactive case-finding generates further health benefits, costing 12,952 GBP/QALY gained. Using ID-LIVER-ML in the proactive-and-reactive population has the highest probability of maximising cost-effectiveness, when valuing QALYs at 20,000 GBP.

**Conclusion:** Smart methods of case-finding and risk-stratification identify people with significant CLD in the community, and are likely to represent good value for money in England.

## 1. INTRODUCTION

It almost seems to go without saying that early detection of chronic liver disease (CLD) will improve patient outcomes and deliver better value to health systems. A substantial number of people live with undiagnosed CLD, which often remains asymptomatic until it becomes life-threatening[1]. Identifying these cases earlier could improve health outcomes and reduce long-term healthcare costs. However, finding people who would benefit from a diagnosis of CLD while making efficient use of NHS resources is challenging. Early detection initiatives have costs and, without targeted approaches, hepatology services risk becoming overwhelmed by referrals of people who ultimately do not have clinically significant disease. While some local commissioners have established referral pathways from primary care, fewer have implemented proactive case-finding strategies for at-risk individuals in the community[2].

Around 90% of CLD mortality in England over the last two decades was lifestyle related, with the most common causes alcohol-related liver disease (ARLD) and metabolic-dysfunction-associated steatotic liver disease (MASLD)[3]. The North West has the highest prevalence of alcohol- and diet-related risk factors for CLD in England[4]. Integrated Diagnostics for Early Detection of Liver Disease (ID-LIVER) was a large, collaborative research project based in Greater Manchester, funded to unite universities, NHS trusts, and small and large commercial partners. It addressed critical gaps around improving the detection of CLD at reversible stages, moving diagnostics and initial management to community-based care, and enabling needs-based diagnostics and interventions[5]. One work-package focused on identifying people in the community at risk of CLD (‘proactive case-finding’), using digital search-tools in routine primary-care data. People with one or more risk factor for ARLD or MASLD were invited to a community liver assessment clinic (CLAC) with a hepatologist and hepatology nurse, held in an accessible location (local healthcare facilities or a mobile screening van)[6]. In another work-package, the investigators used machine-learning algorithms to develop a risk-prediction tool identifying cases with a high probability of significant fibrosis (hereafter ‘ID-LIVER-ML’)[7].

Uniform data were collected for all participants – those in the proactive case-finding population attending a CLAC and routine referrals attending a hepatology outpatient clinic – including liver stiffness measurement (LSM) using vibration-controlled transient elastography. These data were used to train and validate the ID-LIVER-ML model, using LSM ≥ 8.0kPa – indicating high risk of significant fibrosis – as a reference standard.

In this study, we use data from these work-packages to evaluate the cost effectiveness of strategies incorporating one or both of proactive case-finding and enhanced risk-stratification (ID-LIVER-ML), compared with standard referral pathways.

## 2. METHODS

We developed a model-based cost-effectiveness analysis to evaluate the benefits, harms, and costs of different strategies to identifying patients with CLD in the community setting. The analysis conforms to the NICE reference case, adopting an NHS and personal social services perspective on costs[8]. This report follows the Consolidated Health Economic Evaluation Reporting Standards (CHEERS) statement (see *eAppendix 1*)[9].

### 2.1. Strategies evaluated

We evaluate two approaches to case-identification and three approaches to risk-stratification, combining factorially to represent six possible strategies (*Figure 1*).

**Figure 1.**
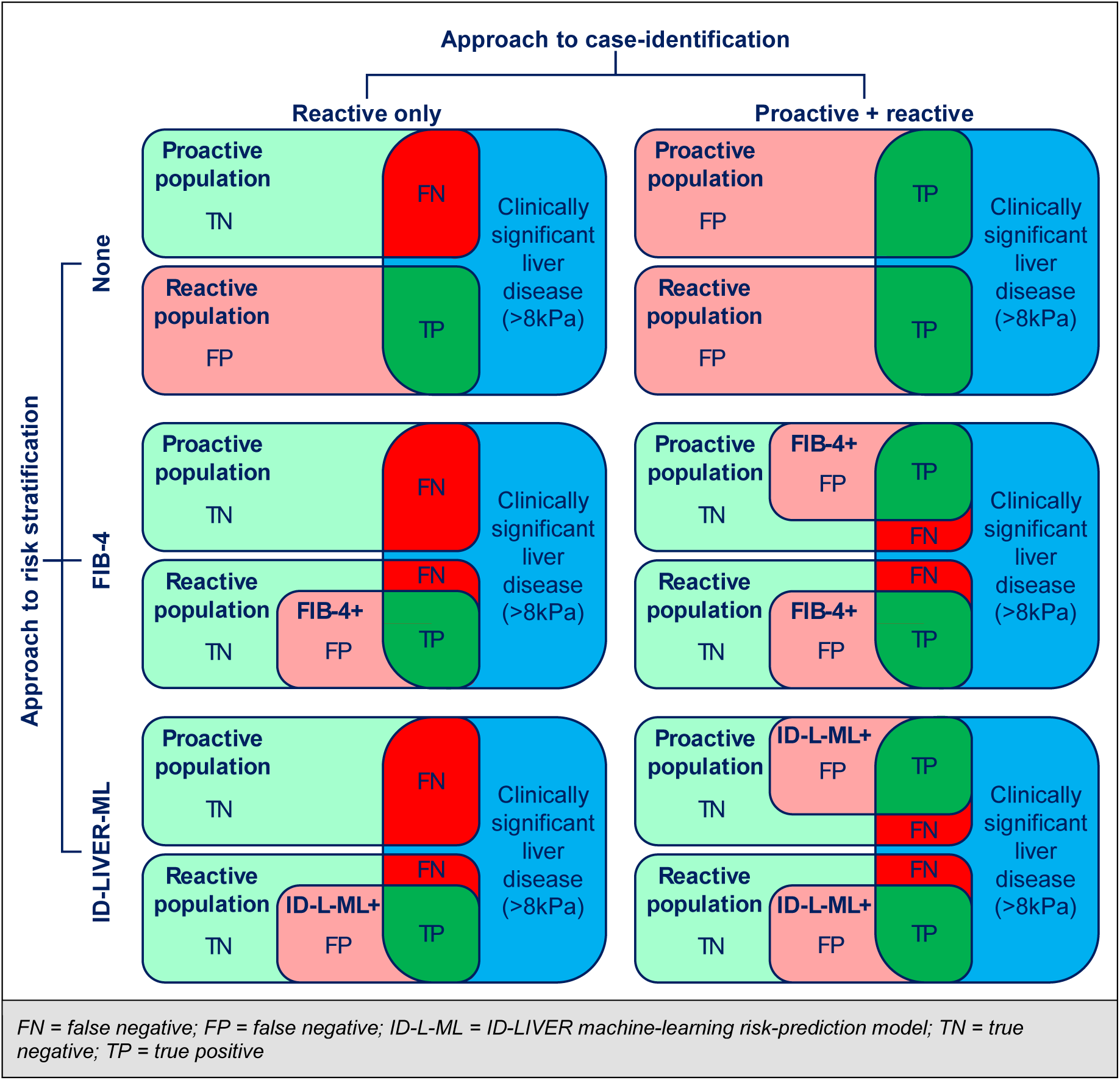
Illustration of the six strategies evaluated, combining two approaches to case-identification and three approaches to risk-stratification

For case-identification, the ‘reactive’ approach represents the current pathway, where hepatology units passively receive referrals, primarily from GPs. The ‘reactive+proactive’ approach supplements routine referrals with case-finding in the community, as explored in ID-LIVER[5]. The three approaches to risk-stratification are: none (everyone referred receives secondary-care review); FIB-4 (limiting secondary-care review to people with a given score); and ID-LIVER-ML (referral only for people with a given probability of significant CLD).

### 2.2. Model structure

We use two independent but structurally identical decision-analytic models – one each for ARLD and MASLD – and weight results together as a final step. The models begin with decision-trees that apportion the cohorts into three categories: minimal fibrosis (equivalent to METAVIR F0/F1), significant fibrosis (F2/F3), and compensated cirrhosis (F4), further subdividing according to the proportion in each category the strategy in question identifies or misidentifies. Then, to simulate the natural and treated history of CLD, and thereby capture the impact of correct identification, we use state-transition (Markov) models; *Figure 2* provides a depiction of their structure. Cohorts enter the state-transition models from the terminal nodes of the decision-tree. Subsequent states reflect long-term consequences of CLD: decompensated cirrhosis (note that this state does not necessarily reflect people experiencing acute decompensating events; rather it represents people who have had at least one such episode), hepatocellular carcinoma (HCC), and death. We distinguish between early HCC (BCLC 0/A), late HCC (BCLC B/C), and HCC arising in a failing liver (BCLC D).

**Figure 2.**
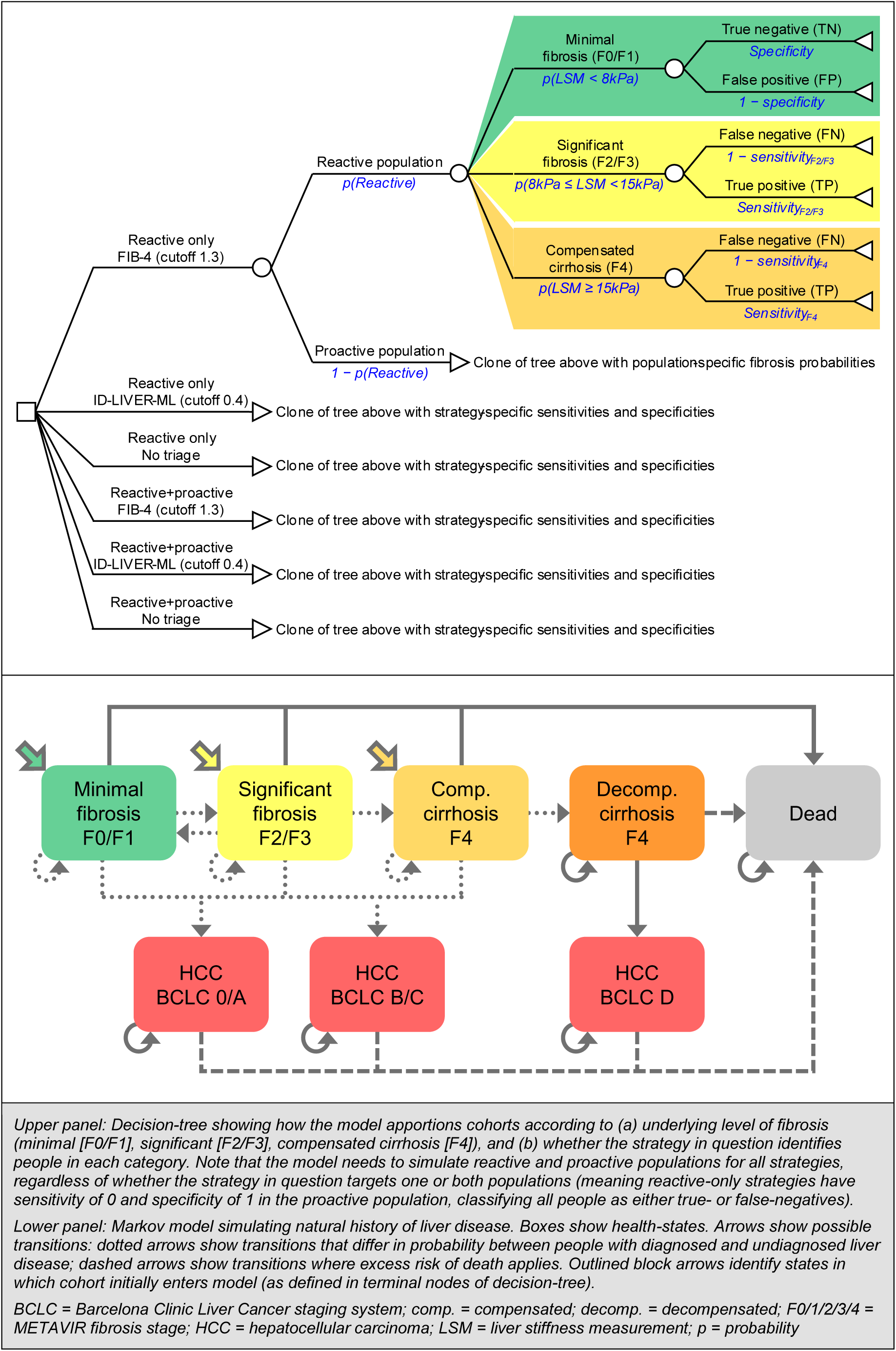
Model structure – decision-tree (identification probabilities) and Markov model (natural history)

Decompensated cirrhosis and HCC are subject to excess, liver-specific mortality; other-cause deaths can occur in any state. Diagnosis of significant fibrosis leads clinicians to recommend lifestyle interventions that may slow disease progression. Thus, people correctly diagnosed have a lower probability of progressing to subsequent states, and may even reverse their liver damage. We assume that regression is not possible from cirrhosis. Once people enter the decompensated cirrhosis state, their CLD becomes known, regardless of whether it was previously diagnosed. People developing HCC come to attention one year later.

We use an annual cycle length with half-cycle correction, and set the model’s time-horizon so that all cohorts reach the age of 100. We apply a discount rate of 3.5% for costs and outcomes measured in quality-adjusted life years (QALYs).

### 2.3. Parameters

#### **2.3.1.** Baseline characteristics of cohorts

We base our simulated populations on data from ID-LIVER, comprising people with ARLD (n=286) and MASLD (n=850) identified via routine referral (n=840) or proactive case-finding (n=296). Starting age and sex depend on aetiology and stage of fibrosis; prevalence of each category of fibrosis depends on aetiology and mode of identification (*Table 1*).

**Table 1.** Summary of model parameters – baseline population

| Parameter | Value (95% CI) | Probabilistic parameters | Source |
| --- | --- | --- | --- |
| Starting age (years) |  |  |  |
| ARLD F0/1 | 55.4 (53.7, 57.1) | Normal: $\mu=55.40$; $\sigma=0.89$ | ID-LIVER cohort |
| ARLD F2/3 | 58.1 (55.4, 60.8) | Normal: $\mu=58.11$; $\sigma=1.37$ | |
| ARLD F4 | 58.6 (55.5, 61.8) | Normal: $\mu=58.63$; $\sigma=1.62$ | |
| MASLD F0/1 | 54.4 (53.3, 55.5) | Normal: $\mu=54.41$; $\sigma=0.55$ | |
| MASLD F2/3 | 57.1 (54.9, 59.2) | Normal: $\mu=57.05$; $\sigma=1.12$ | |
| MASLD F4 | 59.9 (55.8, 64.0) | Normal: $\mu=59.87$; $\sigma=2.09$ | |
| Sex (%male) |  |  |  |
| ARLD F0/1 | 0.665 (0.595, 0.731) | Beta: $\alpha=121$; $\beta=61$ | ID-LIVER cohort |
| ARLD F2/3 | 0.557 (0.432, 0.679) | Beta: $\alpha=34$; $\beta=27$ | |
| ARLD F4 | 0.744 (0.605, 0.861) | Beta: $\alpha=32$; $\beta=11$ | |
| MASLD F0/1 | 0.452 (0.415, 0.489) | Beta: $\alpha=318$; $\beta=386$ | |
| MASLD F2/3 | 0.509 (0.418, 0.599) | Beta: $\alpha=59$; $\beta=57$ | |
| MASLD F4 | 0.600 (0.423, 0.765) | Beta: $\alpha=18$; $\beta=12$ | |
| Proportion of cases reactive |  |  |  |
| ARLD | 0.557 (0.509, 0.604) | Beta: $\alpha=235$; $\beta=187$ | ID-LIVER cohort<br>(financial yr 2022/23) |
| MASLD | 0.631 (0.546, 0.711) | Beta: $\alpha=82$; $\beta=48$ | |
| Proportion of cases with MASLD |  |  |  |
| Reactive | 0.731 (0.700, 0.760) | Beta: $\alpha=614$; $\beta=226$ | ID-LIVER cohort |
| Proactive | 0.797 (0.750, 0.841) | Beta: $\alpha=236$; $\beta=60$ | |
| Weighted average | 0.760 |  |  |
| Proportion with fibrosis at baseline |  |  |  |
| Reactive |  |  |  |
| ARLD |  |  |  |
| F0/1 (LSM < 8kPa) | 0.588 (0.524, 0.652) | Dirichlet:<br>$\alpha_1=133$; $\alpha_2=52$; $\alpha_3=41$ | ID-LIVER cohort |
| F2/3 (8kPa ≤ LSM < 15kPa) | 0.230 (0.178, 0.287) |  |  |
| F4 (15kPa ≤ LSM) | 0.181 (0.134, 0.234) |  |  |
| MASLD |  |  |  |
| F0/1 (LSM < 8kPa) | 0.796 (0.764, 0.827) | Dirichlet:<br>$\alpha_1=489$; $\alpha_2=97$; $\alpha_3=28$ | ID-LIVER cohort |
| F2/3 (8kPa ≤ LSM < 15kPa) | 0.158 (0.130, 0.188) |  |  |
| F4 (15kPa ≤ LSM) | 0.046 (0.031, 0.063) |  |  |
| Proactive |  |  |  |
| ARLD |  |  |  |
| F0/1 (LSM < 8kPa) | 0.817 (0.710, 0.903) | Dirichlet:<br>$\alpha_1=49$; $\alpha_2=9$; $\alpha_3=2$ | ID-LIVER cohort |
| F2/3 (8kPa ≤ LSM < 15kPa) | 0.150 (0.072, 0.250) |  |  |
| F4 (15kPa ≤ LSM) | 0.033 (0.004, 0.091) |  |  |
| MASLD |  |  |  |
| F0/1 (LSM < 8kPa) | 0.911 (0.872, 0.944) | Dirichlet:<br>$\alpha_1=215$; $\alpha_2=19$; $\alpha_3=2$ | ID-LIVER cohort |
| F2/3 (8kPa ≤ LSM < 15kPa) | 0.081 (0.049, 0.118) |  |  |
| F4 (15kPa ≤ LSM) | 0.008 (0.001, 0.023) |  |  |

ARLD = alcohol-related liver disease; F0/1/2/3/4 = METAVIR fibrosis stage; LSM = liver stiffness measurement; MASLD = metabolic-dysfunction-associated steatotic liver disease

**Table 2.**
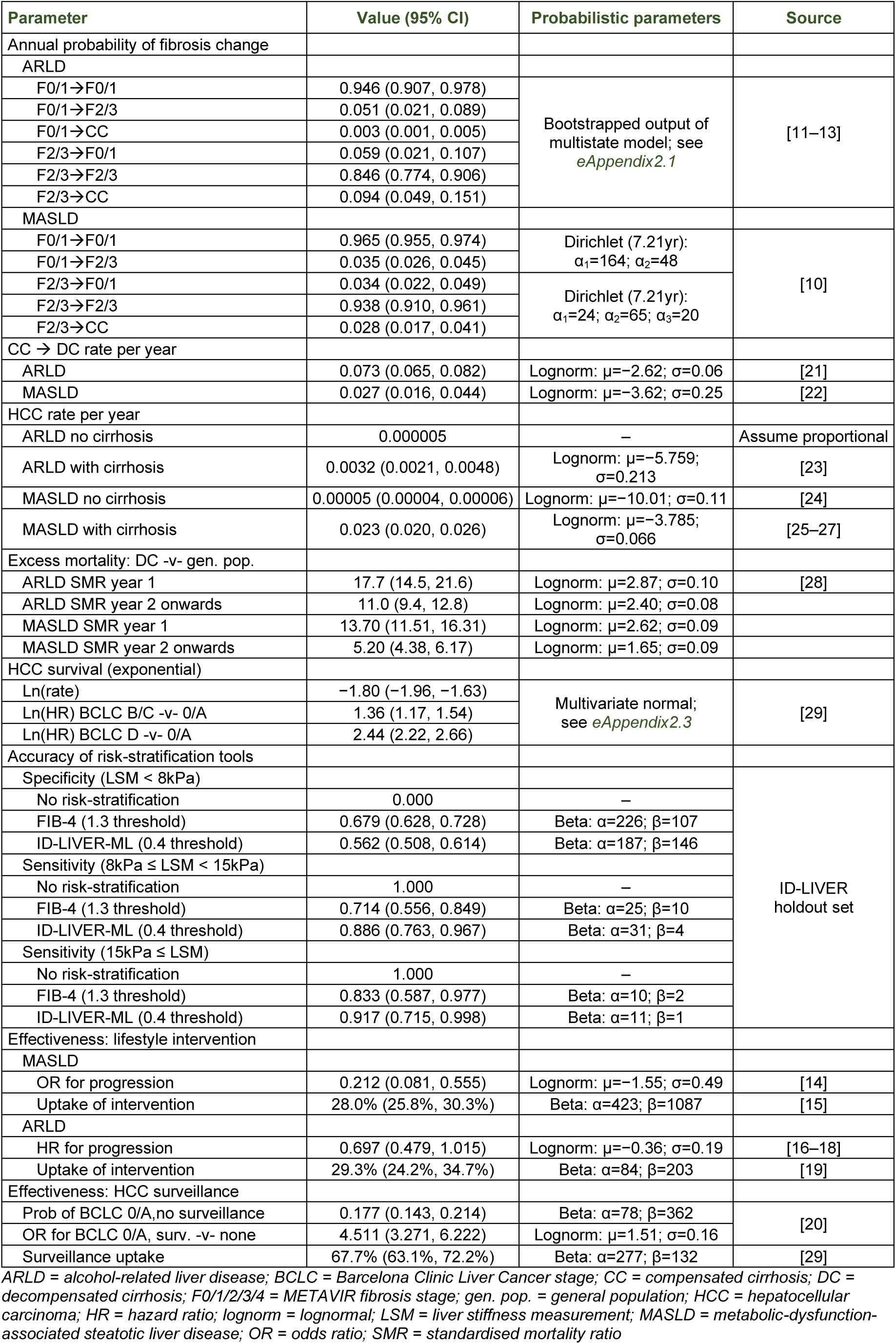
Summary of model parameters – natural history, accuracy of risk-stratification tools, and effectiveness of interventions.

#### **2.3.2.** Natural history of liver disease

To simulate fibrosis progression in MASLD, we use a study reporting sequential biopsies[10]. We derive analogous transition probabilities for ARLD from a multistate model we fitted to patient-level data available in three studies exploring fibrosis progression in liver biopsies11-13](see *eAppendix2.1* for full details).

#### **2.3.3.** Benefits of early detection

Detecting CLD early leads to interventions that could slow or reverse disease progression. For MASLD, we use evidence from a UK study assessing impact of a very-low-calorie diet[14] (*eAppendix2.2* explains parameter derivation). We assume uptake is as observed in a trial assessing a similar intervention in people with diabetes[15]. Effectiveness evidence for ARLD-specific behavioural interventions comes from three retrospective cohort studies reporting association with time to decompensation ; uptake-level comes from a UK-based trial in people with alcohol-use disorder (but not necessarily ARLD)[19]. In our base case, we assume that these effects influence all progression transitions in the model; in a scenario analysis, we explore the impact of limiting the effect to the individual transitions that most closely reflect the effectiveness evidence.

A proportion of people with known compensated cirrhosis also benefits from HCC surveillance. We use UK evidence showing that the probability of finding HCC at an early stage is substantially higher in people undergoing surveillance, and project life expectancy accordingly[20]; *eAppendix2.3* provides details.

#### **2.3.4.** Effectiveness of ID-LIVER proactive case-finding

The effectiveness of ID-LIVER proactive case-finding is a simple function of the proportion of people with significant CLD successfully identified (that is, true-positives with LSM ≥ 8kPa), balanced against the group whose liver health-check did not reveal cause for immediate concern (false-positives). For strategies in which no case-finding takes place, these proportions become false-negatives and true-negatives, respectively.

#### **2.3.5.** Diagnostic accuracy of risk-stratification tools

To estimate the diagnostic accuracy of FIB-4 and ID-LIVER-ML, we use the holdout dataset comprising 380 people from the ID-LIVER cohort whose data were not used in ID-LIVER-ML derivation. We split these individual data into fibrosis categories using LSM as a proxy measure: F0/F1 (LSM < 8.0kPa); F2/F3 (8.0kPa ≤ LSM < 15.0kPa); F4 (15.0kPa ≤ LSM).

We can then observe the number of people in each fibrosis category testing positive at any given threshold of FIB-4 and ID-LIVER-ML, and use these data to define probabilities for use in the decision-tree. For the base case, we set thresholds of 1.3 for FIB-4 (as frequently cited) and 0.4 for ID-LIVER-ML. It is not possible to distinguish ARLD and MASLD in the holdout dataset, so we assume that risk-stratification tools work equally well across both populations.

#### **2.3.6.** Resource-use and costs

Consistent with the study perspective, we account for resource-use associated with diagnosis and liver-related care only. This comprises four main elements: upfront costs (identification, risk-stratification, and initial assessment); ongoing, fibrosis-stage-specific hepatology costs; lifestyle interventions for a proportion of diagnosed people; and HCC treatment. All costs are in 2023/24 British pounds, inflated where necessary using Personal Social Services Research Unit (PSSRU) inflators [32]. Unit-costs mostly derive from NHS Cost Collection and PSSRU. We provide full details in *eAppendix 3*; a summary follows.

We account for primary care appointments and tests for reactive referrals. People assessed in secondary care incur costs of hepatologist time and transient elastography (with nurse time to deliver). We assume hepatology units can deliver these in a ‘one-stop’ clinic, rather than requiring a series of appointments, as we have shown in ID-LIVER[6]. For proactive case-finding, we use estimates of £75 per person for identification (see *eAppendix3.1.2*) and £153 per person for CLAC attendance (*eAppendix3.1.2.2*). We assume that estimating FIB-4 / ID-LIVER-ML incurs no cost (noting that we have already accounted for the blood-tests that underpin them). We explore the possibility that ID-LIVER-ML may ultimately attract a licence-fee in threshold analysis.

For ongoing hepatology care, we only account for liver-related resource-use, so people with undiagnosed disease and those with F0/F1 fibrosis use no healthcare resources. We assume people with diagnosed F2/F3 fibrosis attend one hepatology outpatient appointment per year, including transient elastography and blood-tests. For people with diagnosed compensated cirrhosis and anyone post-decompensation, we rely on costs estimated in NICE guideline NG50 (inflated to 2023/24 values). We add the costs of twice-yearly ultrasound and biochemistry for a proportion of people undergoing HCC surveillance.

For the proportion of people with MASLD accepting lifestyle intervention, we account for dietitian-time and use of meal-replacement products, directly reflecting the very-low-calorie diet we use for effectiveness estimates[14]. For ARLD, we assume a 12-appointment behavioural intervention, as recommended by NICE guideline <u>CG115</u>. See *eAppendix3.3*.

For HCC treatment, we mostly use costs estimated by Cullen *et al*.[34], although a more complex calculation is required for people receiving systemic anticancer therapy; this relies heavily on NICE technology appraisal <u>TA551</u>. *eAppendix3.4* gives details.

#### **2.3.7.** Health-related quality of life

We use EQ-5D-5L measurements from the ID-LIVER cohort, crosswalked to EQ-5D-3L values[35], to estimate health-related quality of life (HRQoL) in people with compensated CLD. The values distinguish between aetiology (ARLD -v-MASLD) and fibrosis stage (F0/F1 -v-F2/F3/F4). We translate absolute estimates into utility multipliers by dividing them by expected HRQoL for age- and sex-matched general population for each category[36]. In our base case, we assume that people retain the HRQoL associated with their baseline fibrosis- state, even if their fibrosis worsens or improves, until they experience decompensation or HCC diagnosis. This is because compensated CLD is largely asymptomatic, and we did not want to assume a causal relationship between liver pathology and HRQoL, when the association between the two is likely mediated by other factors (e.g. comorbidities; extent of obesity / harmful alcohol use). However, the lifestyle interventions that we simulate may also have an impact on these factors, so we explore the impact of allowing HRQoL to vary with fibrosis-state in a scenario analysis.

We calculate a pooled multiplier to reflect progression from compensated to decompensated cirrhosis from studies that report EQ-5D values for both states (*eAppendix4.1*). For HRQoL associated with HCC, we use evidence from a study examining the impact of early- and late-stage HCC diagnosis on HRQoL measured using SF-36[37], which we map to EQ-5D values[38].

### 2.4. Implementation and sensitivity analysis

We built the model in Microsoft Excel. Probabilistic sensitivity analyses rely on 10,000 simulations, sampling all parameters from appropriate uncertainty distributions (mostly empirical; where unavailable, we assume standard error 20% of mean; see *Tables 1–3* and *eAppendices2–3*). We use bootstrapping to account for uncertainty in diagnostic accuracy inputs, taking a random sample of the ID-LIVER holdout population with replacement for each iteration of the model. We performed deterministic (one-way) sensitivity analyses to explore individual parameters’ influence on model outputs, and scenario analyses as noted above (see *eAppendix 9* for a list).

**Table 3.**
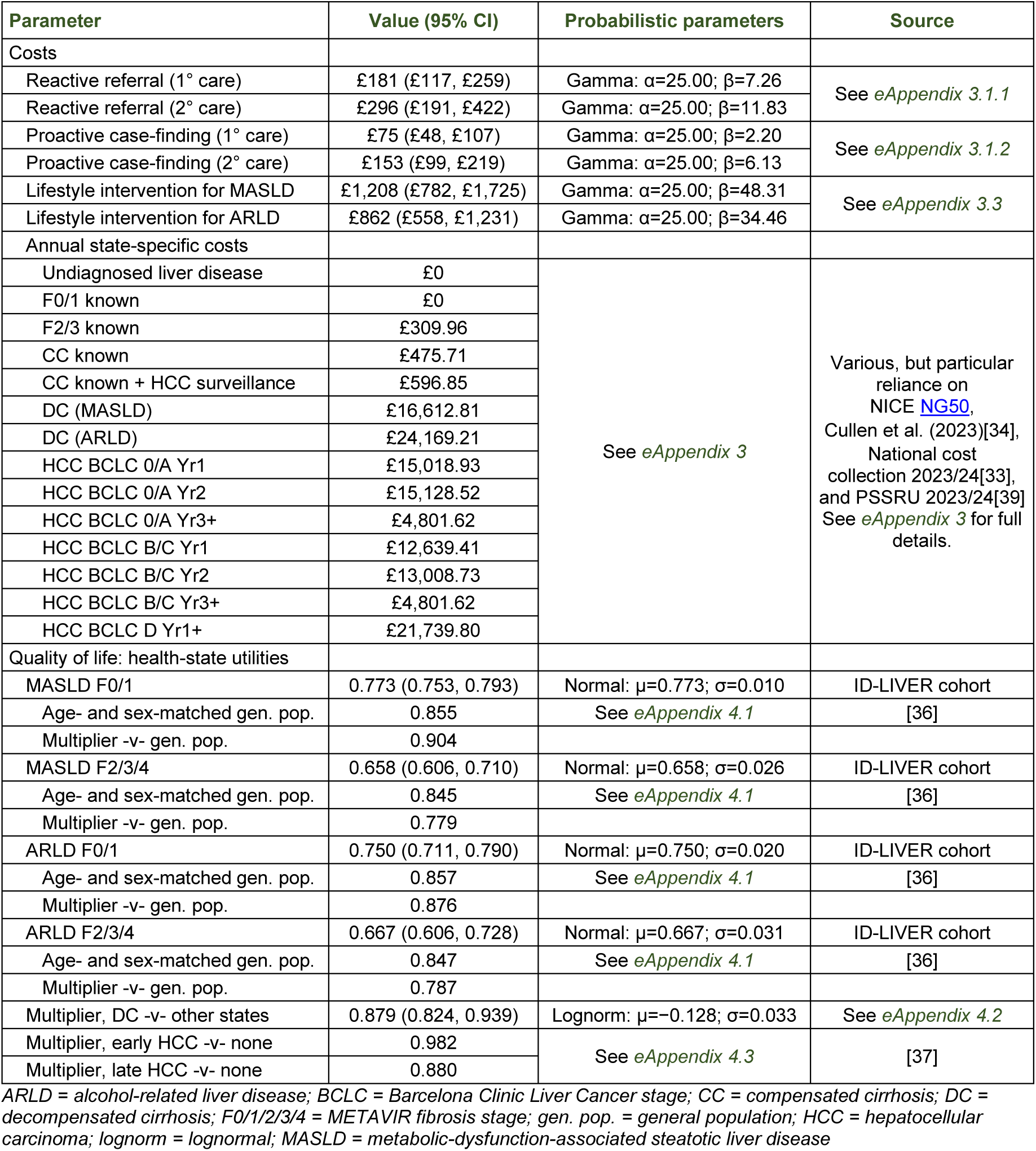
Summary of model parameters – costs and quality of life

### 2.5. Patient and public involvement

ID-LIVER had a collaborative approach to patient and public involvement throughout all stages of project design, development and delivery. This was led by Vocal (wearevocal.org), a not-for-profit health research organisation, and involved regular input from two patient governance advisors with lived experiences of CLD risk factors. They attended work package meetings and participated in a monthly forum with the research team. The team actively consulted underserved community groups with CLD risk factors to improve communication and participation using tailored resources.

## 3. RESULTS

### 3.1. Natural history model

We start by estimating the impact early diagnosis of significant CLD – however achieved – has on expected lifetime costs and QALYs. *Figure 3* shows the difference in prognosis for true-positives and false-negatives in ARLD and MASLD populations (*eAppendix 3* gives separate graphs for each fibrosis state).

**Figure 3.**
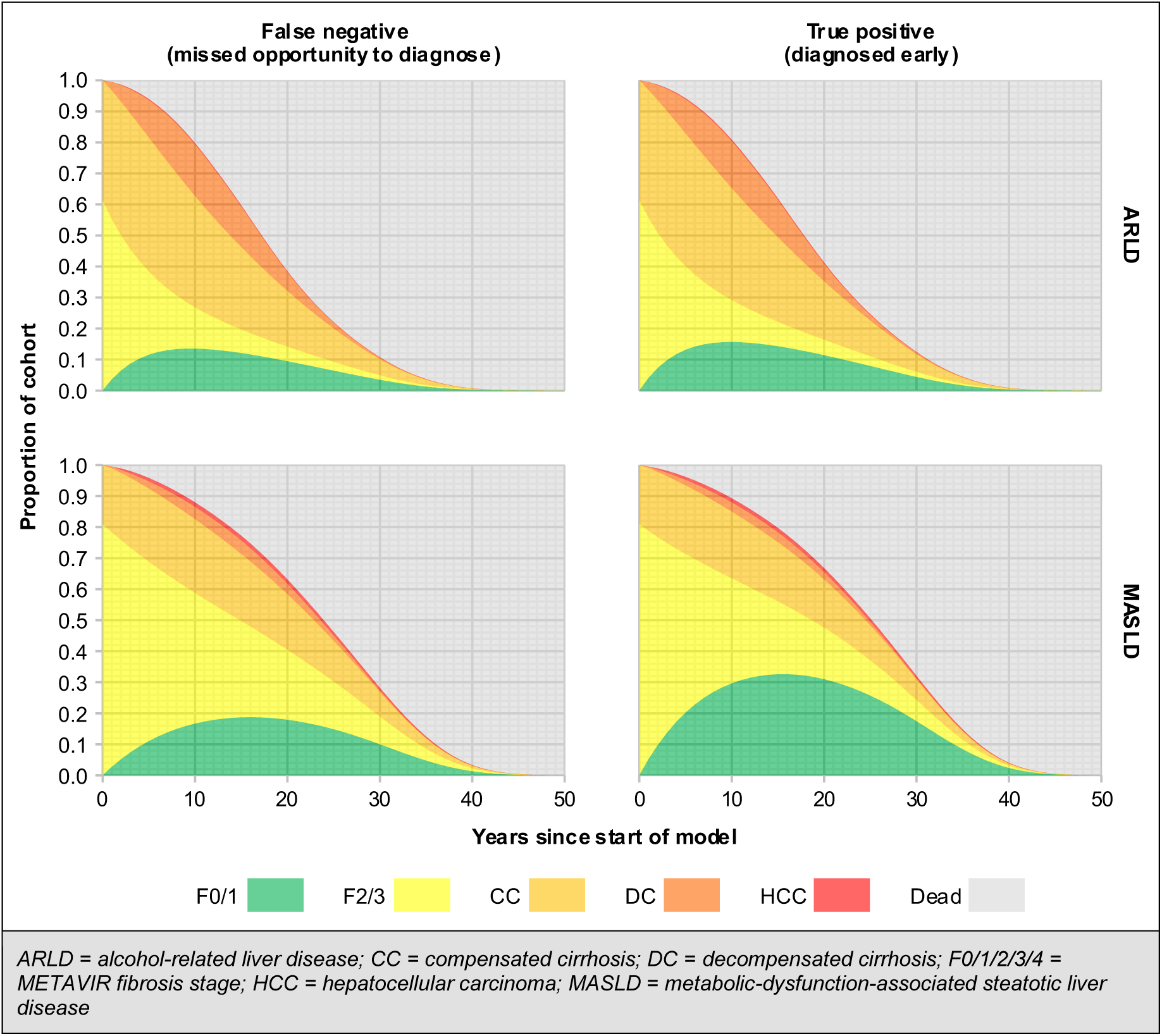
State occupancy over time for people with significant fibrosis, according to aetiology (alcohol-related liver disease-v-metabolic-dysfunction-associated steatotic liver disease) and diagnosis status (false-negative -v-true-positive)

The most obvious impact of successful detection is regression of disease severity from F2/F3 to F0/F1 (i.e. the larger green area in true-positives), particularly in the MASLD population. There is also somewhat lower progression into the compensated cirrhosis, decompensated cirrhosis, and HCC health-states. The HCC health-state is inconspicuous because relatively few people experience this event and most that do move quickly into the dead state. This is particularly true in ARLD, because incidence is lower and mortality associated with decompensated cirrhosis is higher in this population.

By aggregating costs and QALYs associated with predicted time in each state, we can estimate the impact of diagnosing versus not diagnosing CLD (see *Table e10*, in *eAppendix 6*). We find that, if we value QALYs at £20,000 each (NICE-recommended cost-effectiveness threshold), any programme that detects significant CLD at a cost of less than £3,300 per case would generate positive net benefit, compared with no detection. Using these data, we can estimate the cost effectiveness of any risk-stratification strategy for which we know sensitivity, specificity and upfront costs (see *Figure e3*, in *eAppendix 6*).

### 3.2. Cost effectiveness of case-finding and risk-stratification

Figure 4 shows incremental cost-effectiveness results for the six strategies our model simulates. The cheapest strategy is FIB-4-based triage for reactive referrals, because it is the approach that classifies most people – rightly or wrongly – as negative. Switching to risk-stratification with ID-LIVER-ML in the same population identifies more true-positives, generating 0.005 extra QALYs per person at an incremental cost of £52 (∼£10,000 per QALY gained).

**Figure 4.**
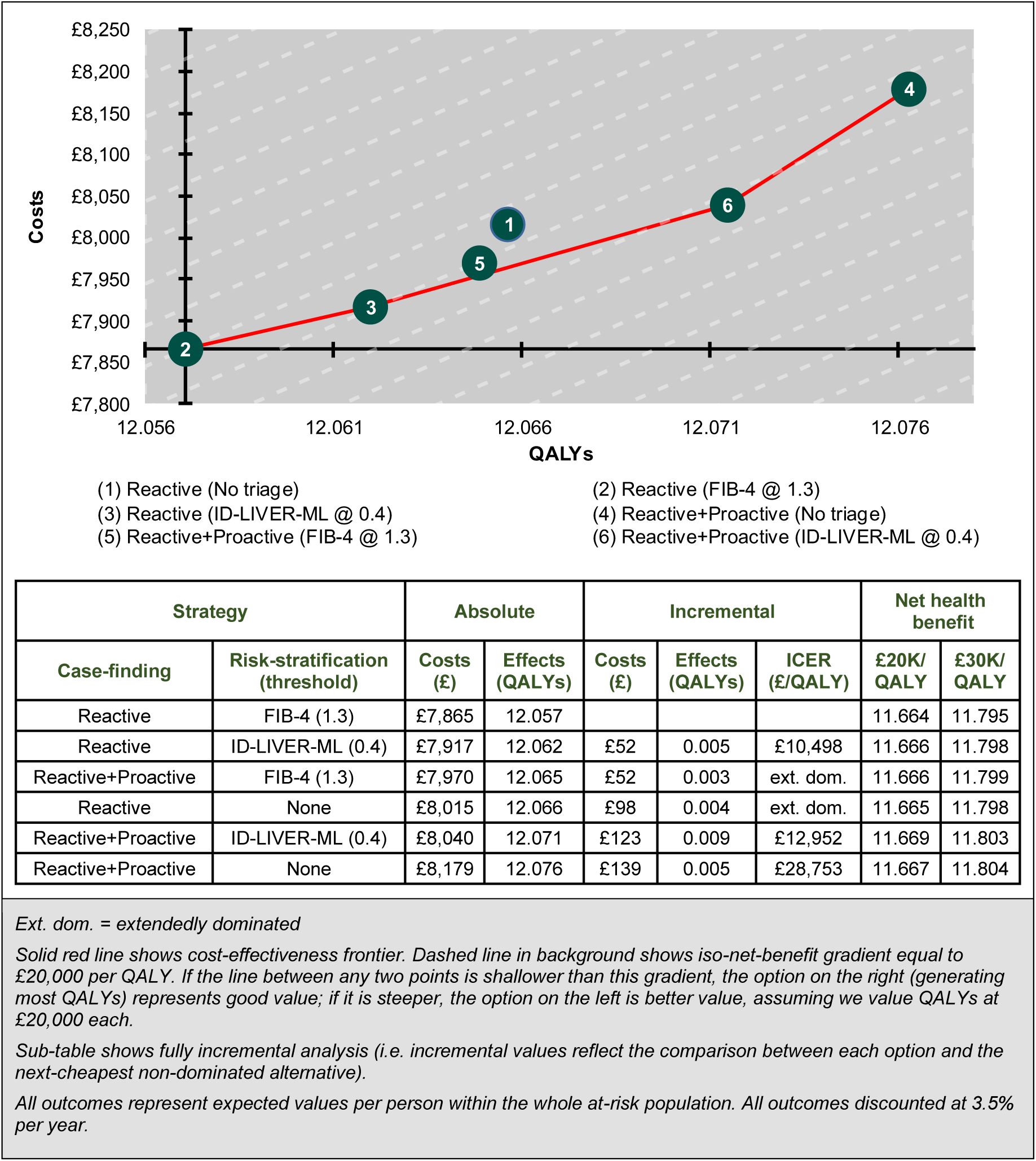
Cost effectiveness of different approaches to case-finding and risk-stratification as assessed in ID-LIVER

Extending the population to include proactive case-finding generates a further 0.009 QALYs, at a cost of £123 per person (∼£13,000 per QALY gained). We would gain most QALYs by combining reactive and proactive populations and applying no risk-stratification (under which strategy, hepatologists will see 100% of people who might be at risk). However, this is also the most expensive approach, and the incremental QALY-gain, compared with ID-LIVER-ML-based triage in the same population, would only be sufficient to counterbalance the incremental cost if we value QALYs at more than £28,000 each.

### 3.3. Sensitivity analyses

Probabilistic sensitivity analysis outputs appear in *eAppendix 7*. If we value QALYs at £20,000 each, the probability that some form of risk-stratification with ID-LIVER-ML is optimal is around 0.7, and the probability that combining it with proactive case-finding provides greatest net benefit is around 0.5. FIB-4-led strategies have less than 0.25 probability of providing best value.

One-way sensitivity analysis (*eAppendix 8*) shows that we are fairly confident that ID-LIVER-ML is a better risk-stratification tool than FIB-4 in the reactive-only population (unless either lifestyle intervention is at the less effective bound of its 95% confidence interval) and, if we extend to the reactive+proactive population, ID-LIVER-ML is better than no risk-stratification. It is somewhat more uncertain whether extending from reactive-only referrals to proactive case-finding represents good value for money; effectiveness estimates for lifestyle interventions are the most obviously influential parameters (explored further in threshold analysis, below).

Scenario analyses (*eAppendix 9*) suggest that attenuating the benefit of diagnosis, by limiting the effect of lifestyle intervention to only one transition, means that the optimal strategy would be to see as few people as possible in secondary care (using FIB-4 in a reactive-only population). Conversely, diagnosis gets more valuable when we assume that HRQoL changes with fibrosis-stage; if we believe this, then it would probably be worth abandoning risk-stratification, and seeing everyone with risk factors in secondary care. When we derive costs of proactive case-finding from the ID-LIVER research budget, overall results do not change materially, though the comparison between ID-LIVER-ML in reactive-only and proactive- and-reactive settings becomes more finely balanced.

Threshold analysis (*eAppendix 10*) shows that lifestyle interventions would only have to be moderately less effective than our base-case estimate to make proactive case-finding bad value (odds ratio ≥0.45 instead of 0.21 for MASLD; hazard ratio ≥0.90 instead of 0.70 for ARLD). When evaluating cutoffs for risk-stratification tests, we find that none generates more net benefit than 0.4 for ID-LIVER-ML. For FIB-4, a cutoff of 0.75 – implying greater sensitivity but worse specificity than current conventions – provides most net benefit, but remains inferior to ID-LIVER-ML at its optimal cutoff. When we apply a cost to the ID-LIVER-ML model, we find that it remains good value for money at any cost ≤£42 per person.

## 4. DISCUSSION

### 4.1. Principal findings

Our study confirms the existence of a reservoir of people with undiagnosed CLD in the community, and shows that we can find some of them in targeted ways that represent good value for money. We also demonstrate that we can improve risk-stratification, both of these proactively identified people and the reactive referrals that remain hepatology services’ stock-in-trade. Although FIB-4-based triage, as commonly used in the UK, is likely to reduce referrals and minimise secondary-care costs, it does so at the expense of ruling out some people with significant CLD. Our analysis suggests that ID-LIVER-ML can minimise such false-negatives, increasing population health at an incremental cost that reflects an effective use of NHS resources.

### 4.2. Strengths

Our study relies on a relatively large dataset, collected in a region with a particular need for diagnostic strategies that will identify people with significant CLD. It is an additional advantage that all participants received a test that is likely to classify their liver health accurately (transient elastography), in contrast to many other studies, in which people who are perceived to be at low risk receive no further investigation. We explicitly simulate the benefits of diagnosis, using best-available empirical data on the healthcare costs, health effects, and likely uptake of lifestyle interventions.

We are able to identify evidence-based, optimal thresholds for risk-stratification tools, based on expected lifetime population-level net benefit, which contrasts with the arbitrary cutoffs that are currently prevalent (e.g. using 1.3 for FIB-4 – which we find is suboptimal – is influenced by a US study that targeted a negative predictive value of 90% without any empirical foundation[31]).

Because of how we structured our analysis, its use is not limited to the approaches we explored in ID-LIVER: we could use it to estimate the cost effectiveness of any case-identification strategy for which we know sensitivity, specificity, and upfront costs.

Our sensitivity analyses enable us to stress-test the analysis and identify areas of greatest uncertainty; unsurprisingly, we find that the parameters with greatest influence on decision-uncertainty are those reflecting the extent to which lifestyle interventions change the prognosis of people whose CLD has been identified.

### 4.3. Limitations

All diagnostic accuracy estimates in our study use vibration-controlled transient elastography as a proxy for true liver pathology. Investigators have shown high concordance between LSM and histology in both ARLD[40-42]and MASLD[43], but it is inevitable that some of the true-positives/false-negatives in our calculations do not have meaningful fibrosis and some of the true-negatives/false-positives do. However, for practical purposes, the distinction is moot: with biopsy becoming ever-rarer, clinicians are seldom directly guided by histology in practice. Moreover, liver biopsy is, itself, an imperfect test that may misclassify people owing to sampling error and/or intra- and inter-observer variability[44]. Therefore, we believe LSM represents as accurate an indicator of fibrosis-stage as is viable in present-day practice.

Our model simulates a one-time opportunity for identification, assuming that any case that is missed will only come to light when symptoms develop. On the one hand, this assumption may overstate value of case-finding, as people may still receive a diagnosis while asymptomatic via later healthcare contacts. On the other, it may understate value of case-finding, as people who are seen but – correctly or incorrectly – discharged may still benefit from lifestyle advice and may also be primed to heed future symptoms.

We have to model the natural history of undiagnosed disease which, by definition, is unobservable. The sequential biopsy studies we use probably provide a reasonable proxy, though any lifestyle modification participants adopted between biopsies may serve to bias true natural progression rates downwards. In this respect, it may not be a bad thing that our ARLD evidence[11-13] comes from a historical era (1970s–80s), during which behaviour-modification interventions were unusual. However, the studies also predate modern-day histopathological criteria, meaning we had to approximate the relationship between reported fibrosis findings and modelled states.

We do not simulate any pharmacological treatments that may slow progression of CLD. NICE’s cirrhosis guideline (NG50) gives a weak (‘consider’) recommendation for beta-blockers for primary prevention of decompensation, based largely on a Spanish randomised trial[45]. Previous cost-effectiveness analyses[46,47]have assumed people with diagnosed CLD gain benefit from pioglitazone; there is some evidence this may be true[48], though we believe it is uncommonly prescribed for this indication in the UK (and is off-label when it is). There is emerging evidence that GLP-1 receptor agonists may reduce LSMs and/or fibrosis . Although none has a marketing authorisation explicitly covering MASLD, NICE currently recommends semaglutide for people with high BMI and ‘at least 1 weight-related comorbidity’ (TA875), as which MASLD would qualify. If any of these agents – or forthcoming MASLD-specific medicines such as resmetirom[51] – provide benefit at acceptable cost, and hepatologists enable access to them for at least some people with significant fibrosis, our analysis will underestimate the value for money with which each true-positive diagnosis is associated.

We chose not to model liver transplantation for people with decompensated cirrhosis (although it is an implicit part of the simulated pathway for early HCC). This outcome is rare for a member of our starting population, and associated with radically different prognosis and costs, making its influence on a decision-problem like this unpredictable.

### 4.4. Comparison with other research

There are three UK-focused analyses assessing referral pathways for people with suspected MASLD. Srivastava *et al*. assessed multiple risk-stratification algorithms relying on noninvasive tests including FIB-4, Enhanced Liver Fibrosis (ELF), and elastography for people in primary care with raised liver enzymes[52]. The authors find that noninvasive tests could increase true-positive referrals to secondary care while reducing false-positives, compared with unassisted GP decision-making (although their estimates of the accuracy of the latter are not based on any empirical data). They also find that identifying people with significant CLD yields net cost-savings, when accounting for prevention of downstream events. This finding contrasts with ours; however, there are multiple differences in methods and inputs. Critically, we specify, evidence, and account for the costs of a mechanism (lifestyle intervention) by which diagnosis may slow disease progression, where Srivastava *et al*. assume benefits without intervention costs. We also note that unit-costs of hepatology appointments have doubled in the time since their analysis. Finally, we note that Srivastava *et al*.’s preferred pathway excludes all people with FIB-4 <1.3; our findings suggest that a nontrivial proportion of such people have significant CLD.

Tanajewski *et al*. model a community risk-stratification pathway using transient elastography to assess people at risk of MASLD[47]. They find a 0.85 probability that the approach is associated with an ICER of £20,000 per QALY or better, compared with standard care. A shortcoming of the model is that it attempts to capture the benefit of diagnosis by assuming 100% of people receive pharmacological treatment with pioglitazone, although this is uncommon in practice. Moreover, the effect-measure comes from a trial using a different drug (rosiglitazone) that has been withdrawn from the UK market (and, in any case, did not show a significant effect in the trial).

Crossan *et al*. assess non-invasive tests to triage secondary-care referrals for people with MASLD[53]. They find that all approaches deliver cost-savings, compared with indiscriminate referrals. They do not model consequences of correct/incorrect diagnoses (e.g. through expected lifetime QALYs). However, they suggest that greatest net benefit, across a range of hypothesised values indicating the relative importance of true- and false-positives, comes from a two-tiered approach using FIB-4 followed by ELF in indeterminate cases.

There are no cost-effectiveness analyses focusing on identifying ARLD in the UK. Asphaug *et al*. simulate various approaches to screening for ARLD in Denmark[54]. They model a hypothetical impact of diagnosis on drinking behaviour, assuming that CLD will only progress in people who continue to drink. They find that risk-stratification with ELF followed by elastography for positive cases is likely to be optimal in primary care, but an elastography-only approach is better in secondary care (although it is unclear why the elastography-only pathway generates fewer QALYs in primary care, when it has most true-positives in both scenarios).

We are not aware of any previous cost-effectiveness analyses looking at proactive case-finding in a broad community at risk for CLD, although one UK-focused cost-effectiveness analysis suggests it would be good value to screen for MASLD in people with type 2 diabetes[46].

### 4.5. Conclusions

Our economic evaluation suggests that early detection of ARLD and MASLD has the anticipated benefits for the person, and can be achieved in ways that represent good value for the system. Our conclusions depend importantly on the effectiveness of interventions to delay progression of CLD. However, any overestimate, in this domain, may be offset by other benefits that we cannot quantify (informal lifestyle advice; possible pharmacological interventions; spillover effects of liver-motivated behaviour-change for nonhepatic outcomes). We continue to undertake proactive case-finding in Greater Manchester, collecting similar data as in ID-LIVER. We intend to use these data to explore potential refinements to the approaches evaluated here. In particular, we aim to improve the specificity of initial digital searches in primary care records, assessing which (combinations of) high-level risk factors identify people at greatest risk of significant CLD.

## Data Availability

All data produced in the present work are contained in the manuscript

## Acknowledgements

We are very grateful for the input of our PPIE governance advisors (Ruth James and Catherine Lennon) and facilitator (Jenny Irvine at Vocal wearevocal.org). Thanks to British Liver Trust for advice on PPIE and subsequent input (Pamela Healy). At Manchester Centre for Health Economics, Anna Donten contributed to costing of the referral pathway and Ali Mahboub-Ahari helped to identify effectiveness and costing evidence for lifestyle interventions. We obtained valuable information to support our micro-costing of proactive case-finding from Northenden Group Practice (practice manager Allison Frith and GP Paul Wright). Mohamed Mostafa and Manish Patel at Jiva.ai generously provided data and insights relating to the ID-LIVER-ML model they developed.

## 5.1. Funding

This work was supported by the UK Research and Innovation [grant numbers UKRI40896 and UKRI10057358].

### 5.2. Declarations of interest

All authors declare they have no competing interests.

### 5.3. Contributions of authors (according to <u>CRediT</u> taxonomy)

## Funding

UK Research and Innovation (UKRI40896 & UKRI10057358).

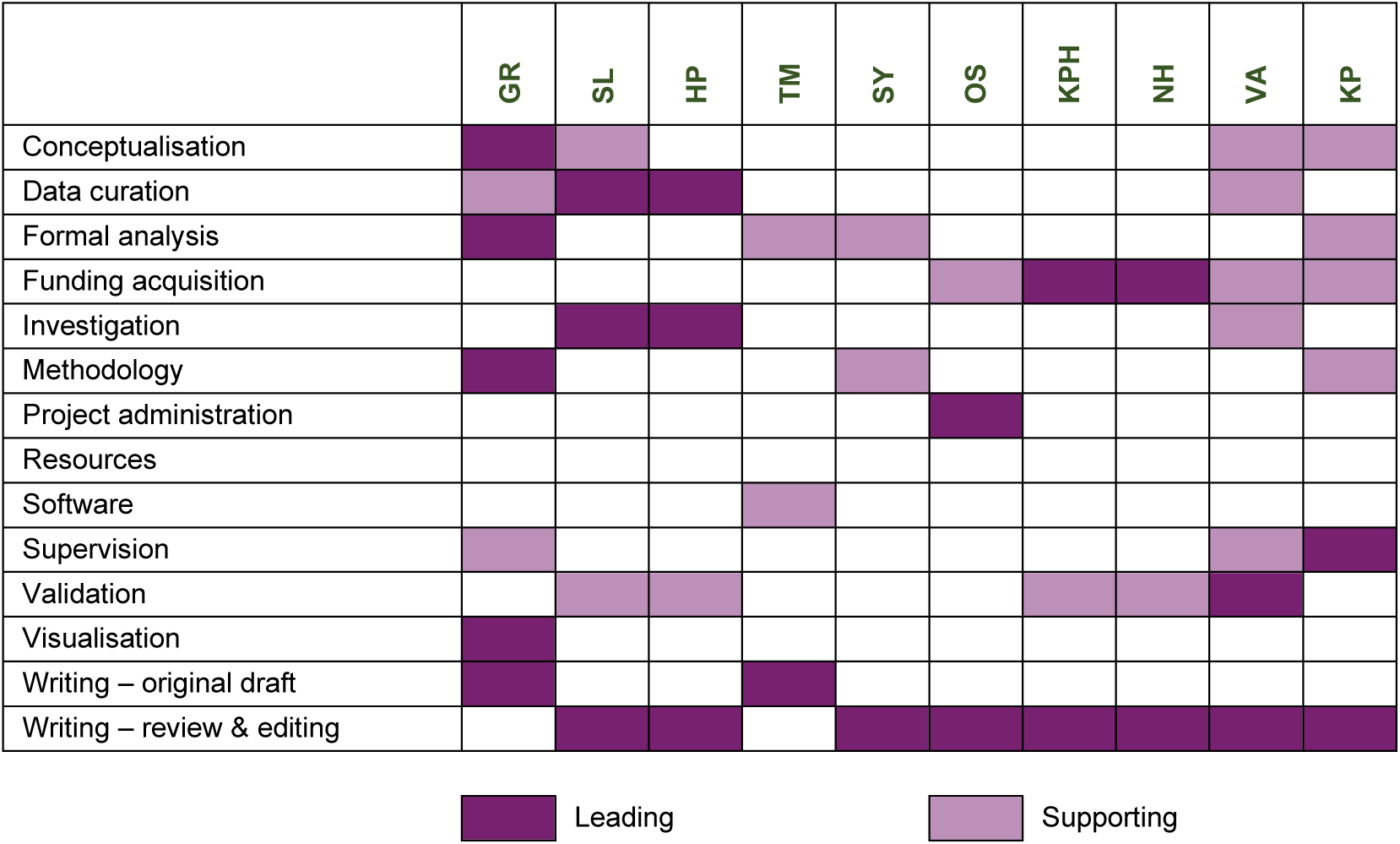

## Notes

### Competing Interest Statement

The authors have declared no competing interest.

### Author Declarations

North of Scotland Research Ethics Committee gave ethical approval for this work (20/NS/0055)

### Summary of Updates

Error in figure 2 corrected (threshold for F4 previously stated 14kPa where it should be 15kPa)

