## Supplementary material for "Proactive case-finding and risk-stratification in people at risk of chronic liver disease in Greater Manchester: a cost-effectiveness analysis"

#### eAppendix 1 CHEERS checklist

| | Item | Guidance for Reporting | Reported on page |
|---|---|---|---|
| **TITLE** | | | |
| Title | 1 | Identify the study as an economic evaluation and specify the interventions being compared. | 1 |
| **ABSTRACT** | | | |
| Abstract | 2 | Provide a structured summary that highlights context, key methods, results and alternative analyses. | 2 |
| **INTRODUCTION** | | | |
| Background and objectives | 3 | Give the context for the study, the study question and its practical relevance for decision making in policy or practice. | 3 |
| **METHODS** | | | |
| Health economic analysis plan | 4 | Indicate whether a health economic analysis plan was developed and where available. | N/A |
| Study population | 5 | Describe characteristics of the study population (such as age range, demographics, socioeconomic, or clinical characteristics). | 8 |
| Setting and location | 6 | Provide relevant contextual information that may influence findings. | 3 |
| Comparators | 7 | Describe the interventions or strategies being compared and why chosen. | 5 |
| Perspective | 8 | State the perspective(s) adopted by the study and why chosen. | 5 |
| Time horizon | 9 | State the time horizon for the study and why appropriate. | 6 |
| Discount rate | 10 | Report the discount rate(s) and reason chosen. | 6 |
| Selection of outcomes | 11 | Describe what outcomes were used as the measure(s) of benefit(s) and harm(s). | 6 |
| Measurement of outcomes | 12 | Describe how outcomes used to capture benefit(s) and harm(s) were measured. | 6 |
| Valuation of outcomes | 13 | Describe the population and methods used to measure and value outcomes. | 9 |
| Measurement and valuation of resources and costs | 14 | Describe how costs were valued. | 11 |
| Currency, price date, and conversion | 15 | Report the dates of the estimated resource quantities and unit costs, plus the currency and year of conversion. | 11 |
| Rationale and description of model | 16 | If modelling is used, describe in detail and why used. Report if the model is publicly available and where it can be accessed. | 6 |
| Analytics and assumptions | 17 | Describe any methods for analysing or statistically transforming data, any extrapolation methods, and approaches for validating any model used. | 11 |
| Characterizing heterogeneity | 18 | Describe any methods used for estimating how the results of the study vary for sub-groups. | 9 |
| Characterizing distributional effects | 19 | Describe how impacts are distributed across different individuals or adjustments made to reflect priority populations. | N/A |
| Characterizing uncertainty | 20 | Describe methods to characterize any sources of uncertainty in the analysis. | 13 |
| Approach to engagement with patients and others affected by the study | 21 | Describe any approaches to engage patients or service recipients, the general public, communities, or stakeholders (e.g., clinicians or payers) in the design of the study. | 14 |
| **RESULTS** | | | |
| Study parameters | 22 | Report all analytic inputs (e.g., values, ranges, references) including uncertainty or distributional assumptions. | 15 |
| Summary of main results | 23 | Report the mean values for the main categories of costs and outcomes of interest and summarise them in the most appropriate overall measure. | 16 |
| Effect of uncertainty | 24 | Describe how uncertainty about analytic judgments, inputs, or projections affect findings. Report the effect of choice of discount rate and time horizon, if applicable. | 17 |
| Effect of engagement with patients and others affected by the study | 25 | Report on any difference patient/service recipient, general public, community, or stakeholder involvement made to the approach or findings of the study | 14 |
| **DISCUSSION** | | | |
| Study findings, limitations, generalizability, and current knowledge | 26 | Report key findings, limitations, ethical or equity considerations not captured, and how these could impact patients, policy, or practice. | 19 |

| | Item | Guidance for Reporting | Reported on page |
|---|---|---|---|
| **OTHER RELEVANT INFORMATION** | | | |
| Source of funding | 27 | Describe how the study was funded and any role of the funder in the identification, design, conduct, and reporting of the analysis | 24 |
| Conflicts of interest | 28 | Report authors conflicts of interest according to journal or International Committee of Medical Journal Editors requirements. | 24 |

*Husereau D, Drummond M, Augustovski F, de Bekker-Grob E, Briggs AH, Carswell C, Caulley L, Chaiyakunapruk N, Greenberg D, Loder E, Mauskopf J, Mullins CD, Petrou S, Pwu RF, Staniszewska S; CHEERS 2022 ISPOR Good Research Practices Task Force. Consolidated Health Economic Evaluation Reporting Standards 2022 (CHEERS 2022) Statement: Updated Reporting Guidance for Health Economic Evaluations. BMJ. 2022;376:e067975.*

*The checklist is Open Access distributed in accordance with the terms of the Creative Commons Attribution (CC BY 4.0) license, which permits others to distribute, remix, adapt and build upon this work, for commercial use, provided the original work is properly cited. See: http://creativecommons.org/licenses/by/4.0/.*

### eAppendix 2 Supplementary modelling methods

#### 2.1. Derivation of transition probabilities for ARLD progression

We derived the transition probabilities for ARLD progression by fitting a multi-state Markov model to patient-level data from three studies examining fibrosis progression in liver biopsies[1–3]. To standardise the histological findings across studies for our model, we mapped the reported fibrosis stages into our three model states, as follows:

- Nakano *et al.* (1982)
  - F0 / F1 : Steatosis
  - F2 / F3 : Steatosis with perivenular fibrosis and fibrosis
  - F4 *:* Incomplete cirrhosis and cirrhosis
- Worner & Lieber (1985)
  - F0 / F1 : Steatosis
  - F2 / F3 : Steatosis with perivenular fibrosis and septal fibrosis
  - F4 *:* Incomplete cirrhosis and cirrhosis
- Marbet *et al.* (1987)
  - F0 / F1 : Fibrosis 0–+
  - F2 / F3 : Fibrosis ++*,* fibrosis +++*, and* early nodular transformation (locally)
  - F4 : Completed nodular transformation (locally)*,* Diffuse early nodular transformation*,* Diffuse early localised established cirrhosis*, and* Diffuse established cirrhosis

We then merged the data into one dataset, which we used to fit the model.

These data represented cross-sectional "snapshots" of patients at various fibrosis stages. We used the {msm} package in R to fit a continuous-time Markov model, assuming homogeneous transition rates between fibrosis states over time.

We modelled disease progression through our three clinically defined states. Given the nature of the data, we specified an initial transition intensity matrix (Q matrix) to reflect plausible transitions between states. The Q matrix allowed for progression from state 1 (F0/F1) to state 2 (F2/F3), bidirectional transitions between states 1 and 2 to capture potential regression and progression, and progression from state 2 to the absorbing state 3 (F4, cirrhosis), with no transitions from state 3.

We estimated the transition intensities using maximum likelihood estimation and calculated bootstrapped confidence intervals to account for parameter uncertainty. From these intensities, we derived the mean transition probability matrices over relevant time intervals to inform our Markov model. We incorporate 10,000 bootstrapped transition probability matrices directly in the decision model, sampling a randomly selected realisation in each iteration of probabilistic analyses.

This approach allowed us to incorporate empirically derived ARLD natural history progression rates into the health economic model.

#### 2.2. Effectiveness of lifestyle intervention for people with MASLD

Our estimate of the effectiveness of lifestyle intervention for people diagnosed with MASLD comes from Scragg *et al.* (2020), a UK before–after study assessing impact of a very-low-calorie diet on weight-loss and liver health[4].

The authors report participants (n=30) had mean LSM of 13.0 (SD 6.6) kPa before intervention, falling to 8.0 (SD 2.9) kPa afterwards. Because 3 people dropped out of the intervention, we adjusted the reported results, assuming that the dropouts would gain no benefit (remain at baseline mean level), resulting in a revised follow-up LSM of 8.5 (SD 3.4) kPa.

To translate these continuous data into the dichotomous effect we need for our model, we use a transformation first derived by Hasselblad and Hedges (1995)[5] and subsequently popularised by Chinn (2000)[6]. First, we calculate a standardised mean difference for the before–after comparison: -0.854 (SE 0.270). Then, on the assumption that the continuous data have an approximately logistic distribution, we can calculate a log-odds ratio using the formula

$$\ln(OR) = SMD\frac{\pi}{\sqrt{3}}$$

This gives us -1.550 (95%CI: -2.510 to -0.589) which, on the natural scale, is an odds ratio of 0.212 (95%CI: 0.081 to 0.555). This implies that people who receive the intervention have 0.2-times the odds of falling above any given LSM cutoff than they did at baseline. We use this odds ratio to adjust transition probabilities between fibrosis states in the model (weighted according to the proportion of people that we assume would accept the intervention – 28.0%, derived from Taylor *et al.*, 2016[7]).

#### 2.3. Effectiveness of surveillance for hepatocellular carcinoma

We use evidence from Haq *et al.*[8] to estimate the impact of HCC surveillance on survival. First, we calculated that people who develop HCC under (fully or partially adherent) surveillance have a 0.493 probability of having an early-stage tumour (stage 0/A according to the Barcelona Clinic Liver Cancer [BCLC] schema). Then, we estimate the extent to which the odds of presenting at the same stage are lower in people who did not receive surveillance (odds ratio = 0.22 [95%CI 0.16 to 0.31]). Combining these two datapoints gives us a probability of 0.177 that people develop HCC at stage 0/A without surveillance. Next, we extracted data from the Kaplan–Meier curves Haq *et al.* provide showing survival according to BCLC stage of HCC at diagnosis, using Guyot *et al.*'s algorithm[9]. We pooled data for BCLC B and C, as these are handled together in our model. Finally, we fitted an exponential survival model to the pseudo-patient-level data we had extracted. *Table e1* shows the fitted coefficients, and provides a variance–covariance matrix, which we used to characterise uncertainty in probabilistic analyses (multivariate normal sampling via Cholesky decomposition).

##### Table e1 Exponential survival model fitted to data extracted from Haq *et al.*[8]

| Term | Estimate (95% confidence interval) | Variance–covariance matrix | | |
|---|---|---|---|---|
| | | Ln(rate) | Ln($HR_{B/C\text{-}v\text{-}0/A}$) | Ln($HR_{D\text{-}v\text{-}0/A}$) |
| Ln(rate) | −1.798 | 0.00714 | −0.00714 | −0.00714 |
| Ln($HR_{B/C\text{-}v\text{-}0/A}$) | 1.356 | −0.00714 | 0.00916 | 0.00714 |
| Ln($HR_{D\text{-}v\text{-}0/A}$) | 2.441 | −0.00714 | 0.00714 | 0.01249 |
| Exponentiated | | | | |
| Rate (BCLC 0/A) | 0.166 (0.140 to 0.196) | | | |
| Hazard ratios | | | | |
| BCLC 0/A | 1 | | | |
| BCLC B/C | 3.88 (3.22 to 4.68) | | | |
| BCLC D | 11.49 (9.23 to 14.30) | | | |

We use an exponential function as it provides a visually adequate fit to the data and, because members of the simulated cohort develop HCC throughout the modelled period, it would not be possible to use a time-varying hazard without substantially increasing model complexity (e.g. with numerous tunnel states or multidimensional transition probability matrices). *Figure e1* illustrates the underlying data and the fitted exponential functions.

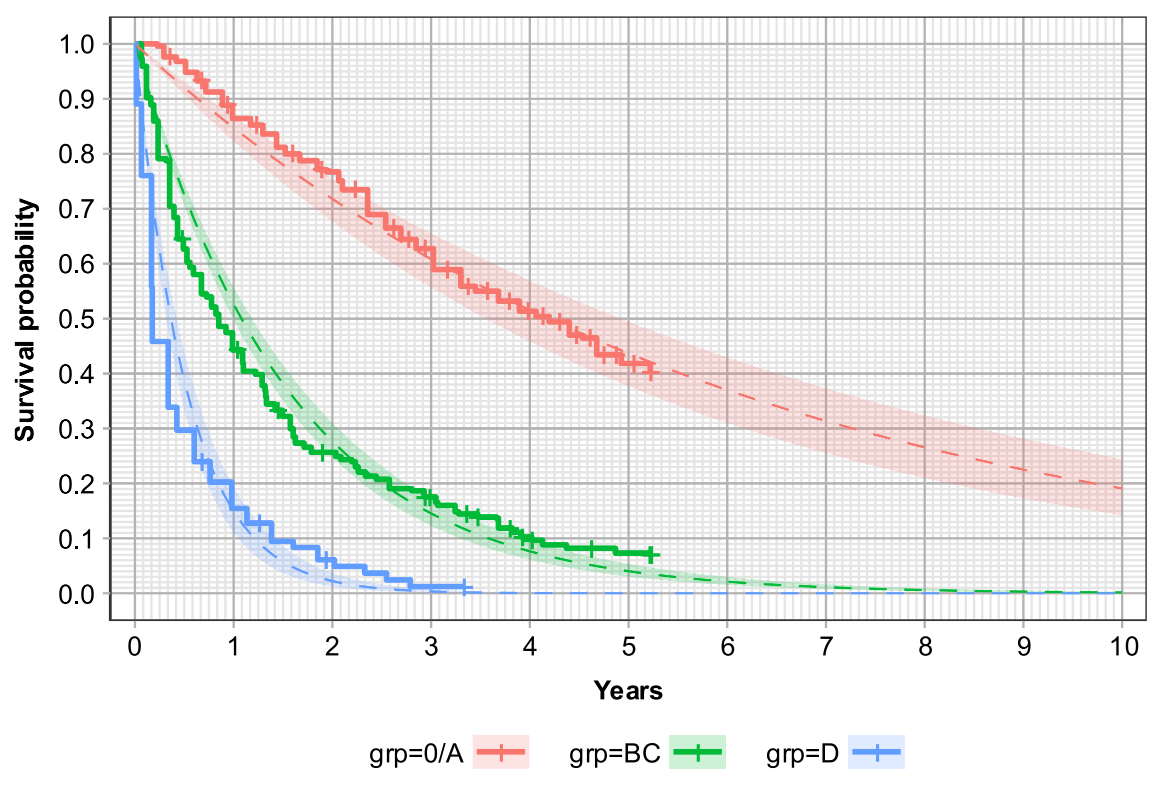

*Solid, stepped lines show empirical survival data (reconstructed from original publication). Dashed, smooth lines show fitted exponential curves. Shaded bands show 95% confidence intervals for fitted curves.*

**Figure e1** **Survival data extracted from Haq *et al.*[8] with overlaid fitted exponential curves**

### eAppendix 3 Cost Inputs

#### 3.1. Upfront costs

##### 3.1.1. Reactive referral

Unit costs for the reactive referral pathway are derived from PSSRU 2023/24 and NHS Cost Collection 2023/24[10,11].

In the reactive referral pathway, we account for primary and secondary care costs. The costs incurred in primary care comprise two GP appointments including a full liver-screen aetiology blood-test and a directly-accessed ultrasound. The unit cost of a standard 10-minute GP appointment is sourced from PSSRU 2023/24[11]. For the cost of the ultrasound, we calculate a weighted average of ultrasound scan costs with a procedural duration above and below 20 minutes. Ultrasound and blood test unit costs are obtained from NHS Cost Collection 2023/24[10]. In total, this amounts to **£181.47**.

Following the results of the diagnostic investigations, patients are initially assessed through a 5-minute consultant-led qualitative triage in secondary care. The hourly cost of a consultant is derived from PSSRU 2023/24[11]. Those stratified to be at risk of significant CLD attend a one-stop outpatient hepatology appointment, including transient elastography. We derive the cost of a first, consultant-led, in-person outpatient appointment from NHS Cost Collection 2023/24[10]. For elastography, we account for equipment costs (sourced from NICE DG48) and 10 minutes of nurse-time (average of band 6 and band 7 pay from PSSRU 2023/24[11])

This amounts to a total of **£295.74**. The whole reactive referral pathway including primary and secondary care costs comes to £477.22.

##### Table e2 Costs associated with reactive referral

| Item | Cost | Notes | Source |
|---|---|---|---|
| Primary care | | | |
| GP appointments | £90.00 | Assume average of 2 appointments before referral | PSSRU 2023/24[11] |
| Liver screen blood tests | £20.49 | Sum of HRG codes: PATH04 (Clinical biochemistry), PATH05 (Haematology), PATH06 (Immunology), PATH07 (Microbiology) | National cost collection 2023/24[10] |
| Ultrasound | £70.98 | Directly accessed ultrasound. Weighted average of RD40Z (duration <20min, without contrast) and RD42Z (duration ≥20min, without contrast) | National cost collection 2023/24[10] |
| Primary care total | £181.47 | | |
| Secondary care | | | |
| Consultant qualitative triage | £8.58 | Assume 5min per case | PSSRU 2023/24[11] |
| Transient elastography | £38.08 | Assumes a 7-year lifetime of a single device, delivering an average of 610 scans per year (average of data from 4 NHS hospitals, including Wythenshawe, Greater Manchester). Accounts for purchase price, 1 additional probe, licence fees for software, service contract, and training costs (all provided to NICE by Echosens). No depreciation. (£28.11; inflated from 2019/20 cost-year) | NICE DG48 |
| Transient elastography nurse time | £11.25 | Assume 10min per case | PSSRU 2023/24[11] |
| Hepatology outpatient appointment | £242.82 | WF01B: Consultant-led, Non-Admitted Face-to-Face Attendance, First | National cost collection 2023/24[10] |
| Secondary care total | £295.74 | | |
| **Whole reactive referral pathway** | **£477.22** | | |

##### 3.1.2. Proactive case-finding

The costs in the proactive referral pathway account for the community-based case-finding efforts which entail using routine primary-care data to identify patients with risk factors for CLD, inviting them to undergo health checks and referring them to the ID-LIVER team.

###### 3.1.2.1. Finding people at risk

###### Current ADA grant (base-case approach)

In the base case of the model, we calculate the cost for proactive case-finding on the basis of our current experience in a successor project to ID-LIVER, the Advanced Diagnostics Accelerator (ADA). Instead of using region-wide electronic searches, as in ID-LIVER, we now work with local GP practices to identify people on their records with one or more risk factor for CLD. Practices are paid a maximum of £5,000 for their participation in ADA (including running searches, sending out invitations by SMS and/or letter, and referring patients who opt in to the CLAC team). To calculate cost per person identified, we simply divide this maximum grant by the average number of people who opt in to attend CLACs per practice. Currently, this figure stands at 67, so we estimate the cost per person identified to be **£74.63**.

We exclude one GP practice (Northenden Group Practice) from this calculation, as ID-LIVER investigators worked closely with practice staff while piloting the approach. This resulted in a high level of buy-in that we accept is unrealistic in routine circumstances. If we were to include Northenden Group Practice in our estimates, the cost per person identified would fall to £47.47.

##### Micro-costing

As part of the ID-LIVER project, we worked directly with GP practices, collecting cost data on the proactive case-finding process. A local GP practice, Northenden Group Practice, provided observed cost data on the admin time to searches, update the website, work on patient invitations, post letters, and pass on details of people who chose to opt in to the CLAC team. This amounts to a total of £1,092. The practice invited 1,000 people, of whom 181 opted in to attend the CLAC. We also account for the cost of GP time to run patient searches for which the reported duration was 30 minutes. To calculate the cost per person identified, we divide the total reported spend by the patient yield at the practice, arriving at a cost of **£6.44**.

**Table e3 Costs associated with finding people at risk of liver disease**

| Item | Value | Notes | Source |
|---|---|---|---|
| GP time to run searches (mins) | 30 | Actioning searches | Personal communication from Northenden Group Practice |
| GP practice admin | £1,092 | Patient invitations, letter postage, website updates. | |
| GP time per hour | £178 | | PSSRU 2023/24[11] |
| Total cost | £1,181 | | |
| Patients seen | 182 | | ID-LIVER |
| **Cost per person identified** | **£6.44** | | |

##### ID-LIVER research costs (scenario analysis)

It is a matter of public record that NorthWest EHealth Limited, the project partner who undertook the electronic searches for ID-LIVER, received £50,982 for their contribution to the research project. Given the final yield of 296 people attending CLACs during the funded project, this amounts to a cost per person identified of **£172.24**. We explore the impact of using this cost in a scenario analysis, as it factually represents the expenditure in the project. However, we have subsequently demonstrated that, by working directly with GP practices, we can achieve the same thing at substantially lower cost (see above), so we take the view that the research cost is not representative of expected expenditure in practice.

##### 3.1.2.2. Community liver assessment clinic

###### Micro-costing

In the micro-costing exercise for the community liver assessment clinics (CLACs), we calculate the costs of assessing patients referred to a CLAC following identification. These costs account for the clinic location which either entails the use of local healthcare facilities or a mobile screening van. In ID-LIVER, a mobile screening van was used at 4 clinic sites, over 28 days in total across all sites, during which 238 patients were seen. The costs include a one-off set-up and admin fee per site, and a daily van rate. Alternatively, the costing for CLACs using local GP facilities account for 9 days of room hire, and 2 admin staff working 7.5 hours daily. These costs are based on observed data from an ID-LIVER GP site where 182 patients attended a CLAC.

For both clinic locations, we account for staff time for a hepatologist (registrar) and hepatology nurse based on a 30-minute appointment duration during which patients receive a blood test and undergo transient elastography. Staff-time unit-costs for a hepatology registrar and nurse (average of band 6 and band 7 pay) come from PSSRU 2023/24[11]. We source the costs of a liver screen blood test from NHS Cost Collection 2023/24[10] and transient elastography equipment from NICE DG48. The appointment duration, room hire, admin time, and mobile van screening costs represent real-world estimates based on observations from the ID-LIVER project. Considering the patient yield for the mobile screening vans and local GP CLACs, the estimated cost per person amounts to **£153.33** and **£146.60** respectively. *Table e4* provides details.

#### Table e4 Costs associated with community liver assessment clinics

| Item | Cost | Notes | Source |
|---|---|---|---|
| Staff time and tests per person seen | | | |
| Hepatology registrar time (hours) | 0.5 | | Authors' assumptions, based on experience in ID-LIVER |
| Nurse time (hours) | 0.5 | In theory, nurses can deliver elastography in less time than this (we estimate a usual duration of 10 minutes). However, we find that, in space-restricted settings, it is not possible to run multiple activities concurrently while maintaining patient confidentiality. Therefore, we make the assumption that both the hepatologist and the nurse are effectively committed to a single patient for the duration of each 30-minute appointment. | |
| Hepatology registrar per hour | £79.00 | | PSSRU 2023/24[11] |
| Nurse per hour | £67.50 | Average of band 6 and band 7 | |
| Transient elastography equipment costs | | | |
| Pay-per-scan business model | £68.26 | Provided to NICE by Echosens (£58; inflated from 2019/20 cost-year) | NICE DG48 |
| Outright purchase | £33.08 | Assumes a 7-year lifetime of a single device, delivering an average of 610 scans per year (average of data from 4 NHS hospitals, including Wythenshawe, Greater Manchester). Accounts for purchase price, 1 additional probe, licence fees for software, service contract, and training costs (all provided to NICE by Echosens). No depreciation. (£28.11; inflated from 2019/20 cost-year) | |
| Liver screen blood test | £20.49 | Sum of HRG codes: PATH04 (Clinical biochemistry), PATH05 (Haematology), PATH06 (Immunology), PATH07 (Microbiology) | National cost collection 2023/24[10] |
| Total staff time and tests | | | |
| Assuming pay-per-scan elastography | £162.00 | | |
| Assuming outright purchase | £126.82 | | |
| Consultation facilities | | | |
| Option 1: mobile screening van | | | |
| Set-up fee per site | £256 | Driver time, set-up/close-down, training | Personal communication, Manchester University NHS Foundation Trust Research & Innovation |
| Admin fee per site | £250 | Admin set-up process | |
| Daily van rate | £153 | Running costs, petrol, equipment usage | |
| Number of clinic sites | 4 | | ID-LIVER |
| Total number of days | 28 | | |
| Total van costs | £6,308 | | |
| Total number of patients seen | 238 | | |
| Cost per person (screening van) | £26.50 | | |
| Option 2: room hire | | | |
| Daily room hire | £175 | Paid daily | Personal communication from Northenden Group Practice |
| Reception / admin staff per day | £225 | 2 admin staff at £15 per hour for 7.5 hours | |
| Number of days | 9 | | ID-LIVER |
| Total room hire / admin staff cost | £3,600 | | |
| Total number of patients seen | 181 | | |
| Cost per person (room-hire) | £19.89 | | |
| **Total cost per person** | | | |
| **Base case** | **£153.33** | Assumes outright purchase of transient elastography equipment and mobile screening van consultation costs | |
| Most expensive scenario | £188.51 | Pay-per-scan transient elastography equipment and mobile screening van consultation costs | |

##### National HRGs

As a simple alternative to our micro-costing, we also explored using national HRGs for a hepatology appointment and transient elastography costs (i.e. exactly the same as specified for the secondary care component of the reactive referral pathway excluding the consultant-led qualitative triage). This amounts to £287.16 per person seen.

#### 3.2. Ongoing fibrosis-stage-specific hepatology costs

Patients diagnosed with F2/3 fibrosis are referred to secondary care. The costs for this health state are estimated to be a total of **£309.96** per year. This accounts for a single hepatology outpatient appointment per year during which patients receive a full liver screen blood test and transient elastography. The costs of the appointment, blood test, and transient elastography are derived from NHS Cost Collection 2023/24[10].

We derive costs for cirrhosis from NICE guideline NG50. We inflate the 6-monthly compensated and decompensated cirrhosis health-state costs for MASLD and ARLD to 2023/24 prices and double them to arrive at the equivalent cost per yearly cycle. For compensated cirrhosis, these costs comprise of a hepatologist appointment and a combination of tests including a full blood count, international normalized ratio and liver blood test. This amounts to **£475.71** for both ARLD and MASLD patients. Costs for decompensated cirrhosis account for three hepatologist appointments, the same combination of tests and include complication costs. In total, this amounts to **£24,169.21** for ARLD patients and **£16,612.81** for MASLD patients. As we only account for liver-related resource use, people in the F0/F1 health states and those who are undiagnosed incur no costs.

##### Table e5 Costs of ongoing hepatology care

| Health state | Cost | Notes | Source |
|---|---|---|---|
| MASLD | | | |
| F0/1 | £0.00 | | |
| F2/3 | £309.96 | 1 hepatologist outpatient appointment, full liver screen blood test, and transient elastography. | NHS Cost Collection 2023/24[10] |
| Compensated cirrhosis | £475.71 | 1 hepatologist appointment, full blood count, international normalized ratio test, and liver blood test. Inflated from 2013/14 to 2023/24 prices and doubled (NICE model has 6-month cycle where ours in 12-months).<br>Surveillance for HCC added separately for a proportion of people; see below. | NICE guideline NG50 |
| Decompensated cirrhosis | £16,612.81 | 3 hepatologist appointments, full blood count test, international normalized ratio test, liver blood test, and complication costs. Inflated from 2013/14 to 2023/24 prices and doubled (NICE model has 6-month cycle where ours in 12-months). | NICE guideline NG50 |
| ARLD | | | |
| F0/1 | £0.00 | | |
| F2/3 | £309.96 | 1 hepatologist outpatient appointment, full liver screen blood test, and transient elastography. | NHS Cost Collection 2023/24[10] |
| Compensated cirrhosis | £475.71 | 1 hepatologist appointment, a full blood count test, international normalized ratio test, and liver blood test. Inflated from 2013/14 to 2023/24 prices and doubled (NICE model has 6-month cycle where ours in 12-months). | NICE guideline NG50 |
| Decompensated cirrhosis | £24,169.21 | 3 hepatologist appointments, a full blood count test, international normalized ratio test, liver blood test, and 50% increased complication costs. Inflated from 2013/14 to 2023/24 prices and doubled (NICE model has 6-month cycle where ours in 12-months). | NICE guideline NG50 |

*ARLD = alcohol-related liver disease; F0/1/2/3/4 = METAVIR fibrosis stage; MASLD = metabolic-dysfunction-associated steatotic liver disease*

#### 3.2.1. Surveillance for HCC

For the patients undergoing HCC surveillance, we account for the cost of each visit. Visits comprise of a clinical biochemistry blood test and a directly accessed ultrasound. These unit costs are both sourced from NHS Cost Collection 23/24[10]. The visit frequency is determined by the level of adherence, with two visits yearly for adherent patients and one for non-adherent patients. The probability of adherence is estimated to be 77% as derived from Haq et al.[8] which looked at adherence to HCC surveillance and its impact on survival in a UK mixed aetiology cirrhosis population. The estimated total cost per year is £121.14.

##### Table e6 Costs associated with surveillance for HCC

| Item | Cost | Notes | Source |
|---|---|---|---|
| Clinical biochemistry blood test | £1.53 | | NHS Cost Collection 2023/24[10] |
| Outpatient ultrasound scan | £67.05 | Weighted average of ultrasound procedure costs lasting over and below 20 minutes. | NHS Cost Collection 2023/24[10] |
| Cost per visit | £68.58 | | |
| Total cost per year | £121.14 | Calculated based on a 77% probability of adherence (Haq et al., 2021), with adherent patients attending two appointments and non-adherent patients attending one. | |

#### 3.3. Lifestyle intervention costs

Patients identified to have MASLD with significant or more severe fibrosis are referred to a lifestyle intervention in form of a dietary intervention. Depending on the uptake level, the incurred costs account for the very low-calorie diet (VLCD) offered to patients. We calculate a mean price per serving using information on the prices and servings of the recommended meal replacement products. Unit costs for these products are obtained from Optifast[12]. Using assumptions about the servings per week derived from Scragg et al.[4], we subsequently estimate the total price of the diet. Additionally, we account for staff time for a band 7 Dietitian per NICE guideline CG189, assuming patients also attend 12 dietitian appointments each for the duration of 30 minutes. Staff costs are obtained from PSSRU 2023/24[11], and assumptions about the number of and duration of the appointments from NICE guideline CG189. The total intervention cost amounts to £1,207.69, which gives an average cost of £416.19 per diagnosed person based on our estimate that 28.0% of people will choose to accept the intervention[13].

For ARLD patients, we account for staff time for a band 7 clinical psychologist, assuming patients who accept treatment undergo a 12-appointment behavioural intervention, each for the duration of an hour. Staff costs are also obtained from PSSRU 2023/24[11] with assumptions about appointment frequency and duration sourced from NICE CG115. This adds up to a total intervention cost of £861.54 resulting in an average cost of £252.16 based on a 29.3% uptake level[14].

##### Table e7 Costs associated with lifestyle interventions

| Item | Cost | Notes | Source |
|---|---|---|---|
| MASLD (very low-calorie diet) | | | |
| Meal replacement products (per serving) | £2.69 | Calculated by dividing the price per pack of each Optifast meal replacement product by the servings per pack | Optifast website |
| Total price | £791.49 | 294 servings over 12 weeks | Scragg *et al.* (2021) |
| Dietitian cost | £416.19 | 12 × 30-minute appointments with band 7 dietitian | PSSRU 2023/24[11]<br>NICE CG189 |
| Total intervention cost | £1,207.69 | | |
| Mean intervention cost per diagnosed person | £338.31 | Assumes 28.0% uptake | [7] |
| ARLD (behavioural intervention) | | | |
| Total intervention cost | £861.54 | 12 × 1-hour appointments with band 7 clinical psychologist | PSSRU 2023/24[11]<br>NICE CG115 |
| Mean intervention cost per diagnosed person | £252.16 | Assumes 29.3% uptake | [14] |

*ARLD = alcohol-related liver disease; MASLD = metabolic-dysfunction-associated steatotic liver disease*

#### 3.4. HCC treatment

##### Table e8 Costs associated with treatment for HCC

| Item | Costs | Notes | Source |
|---|---|---|---|
| Compensated cirrhosis | | | |
| Ablation and curative | £13,759 | Obtained 2-year mean cost estimates from authors of Cullen et al. (2023), inflated these values from 2018/19 to 2023/24 prices and divided by two to derive the per-cycle treatment cost. In probabilistic analysis, we assume SE is 20% of mean, in absence of empirical estimates of uncertainty. | Cullen et al. (2023)[15] |
| Ablation and palliative | £18,405 | | |
| Initial palliative and curative | £27,851 | | |
| Liver resection and other | £11,637 | | |
| Liver transplant and other | £37,910 | | |
| Cytotoxic chemotherapy and other | £12,739 | | |
| Other palliative and other | £9,794 | | |
| TACE and other | £15,230 | | |
| No active treatment | £4,802 | | |
| Decompensated cirrhosis | | | |
| Initial palliative and curative | £35,779 | | |
| Other palliative and other | £11,686 | | |
| No active treatment | £4,832 | | |
| Systemic therapy | | | |
| Progression free | £28,498.12 | Based on NICE appraisals, unit costs updated or values inflated | NICE TA474 |
| Post-progression | £37,562.72 | | NICE TA551 |
| Proportion of people receiving each treatment | | | |
| TNM stage I (~= BCLC 0/A) | | Probabilistic parameters: Dirichlet ($\alpha_1$=229, $\alpha_2$=25, $\alpha_3$=157, $\alpha_4$=164, $\alpha_5$=208, $\alpha_6$=82) | Driver (2022)[16] |
| Best supportive care | 0.2647 | | |
| Sorafenib | 0.0289 | | |
| TACE | 0.1815 | | |
| Ablation | 0.1896 | | |
| Resection | 0.2405 | | |
| Transplantation | 0.0948 | | |
| TNM stage II-IV (~= BCLC B/C) | | Probabilistic parameters: Dirichlet ($\alpha_1$=2,523, $\alpha_2$=385, $\alpha_3$=487, $\alpha_4$=132, $\alpha_5$=323, $\alpha_6$=103) | |
| Best supportive care | 0.6382 | | |
| Sorafenib | 0.0974 | | |
| TACE | 0.1232 | | |
| Ablation | 0.0334 | | |
| Resection | 0.0817 | | |
| Transplantation | 0.0261 | | |
| BCLC D | | | |
| Best supportive care | 1.0000 | Assumption | |
| Costs per person per year | | | |
| BCLC 0/A | | | |
| Year 1 | £15,018.93 | Weighted averages of the above | |
| Year 2 | £15,128.52 | | |
| Year 3 onwards | £4,801.62 | Assume as for no active treatment | |
| BCLC B/C | | | |
| Year 1 | £12,639.41 | Weighted averages of the above | |
| Year 2 | £13,008.73 | | |
| Year 3 onwards | £4,801.62 | Assume as for no active treatment | |
| BCLC D | | | |
| Year 1 onwards | £21,739.80 | BSC costs plus post-decompensation costs (see *Table e5*) | |

*BCLC = Barcelona Clinic Liver Cancer stage; BSC = best supportive care; TACE = transarterial chemoembolization; TNM = Tumour–Node–Metastasis classification of malignant tumours*

### eAppendix 4 Health-related quality of life inputs

#### 4.1. No fibrosis or compensated fibrosis

As well as dividing according to aetiology of liver disease (ARLD -v- MASLD), we split HRQoL values derived from the ID-LIVER cohort according to level of fibrosis (F0/F1 -v- F2/F3/F4). We explored introducing an additional distinction between F2/F3 disease and compensated cirrhosis (F4), in line with the states in our natural history model. However, there was no evidence of difference in EQ-5D-3L utility between people with F2/F3 disease and those with F4; this is consistent with the observation that compensated CLD is largely asymptomatic.

We compared observed EQ-5D-3L utility with expected utility for the general population as reported by Hernández Alava *et al.*[17], age- and sex-matched for each category. Dividing the former by the latter provides a multiplier that we apply in the model to expected utility for the cohort as it ages, which enables us to account for decreasing HRQoL with age. In probabilistic analyses, we incorporate uncertainty in the estimates of general population utility by sampling coefficients for Hernández Alava *et al.*'s model from a multivariate normal distribution, using the variance–covariance matrix the authors provide. We then recalculate the relevant multipliers for each iteration of the probabilistic model.

#### 4.2. Decompensated cirrhosis

*Table e9* shows how we estimate health-related quality of life (utility) for people with decompensated cirrhosis. By pooling the ratio of means in 10 studies reporting EQ-5D utilities for populations with compensated and decompensated cirrhosis, we estimate a multiplier, which we then apply to the relevant age- and sex-adjusted utility for people with F2/F3/F4 fibrosis (see main text).

##### Table e9 Health-related quality of life associated with decompensated cirrhosis compared with compensated cirrhosis

| Study | Country | Population | Compensated | | Decompensated | | Multiplier (ratio of means) |
|---|---|---|---|---|---|---|---|
| | | | N | Utility (95%CI) | N | Utility (95%CI) | |
| Bjornsson et al., 2009[18] | Sweden | HCV | 76 | 0.749 (0.700 to 0.795) | 53 | 0.656 (0.583 to 0.726) | 0.876 (0.772 to 0.994) |
| Chong et al., 2003[19] | Canada | HCV | 24 | 0.750 (0.660 to 0.830) | 9 | 0.660 (0.448 to 0.843) | 0.880 (0.637 to 1.216) |
| Cortesi et al., 2020[20] | Italy | CLD | 574 | 0.891 (0.881 to 0.901) | 523 | 0.859 (0.847 to 0.871) | 0.964 (0.947 to 0.981) |
| Pol et al., 2015[21] | France, Germany, UK | HCV | 101 | 0.670 (0.610 to 0.727) | 25 | 0.510 (0.373 to 0.646) | 0.761 (0.574 to 1.010) |
| Samp et al., 2015[22] | France | HCV | 18 | 0.622 (0.532 to 0.708) | 11 | 0.405 (0.288 to 0.528) | 0.651 (0.468 to 0.905) |
| Sugimori et al., 2022[23] | Japan | HBV | 141 | 0.845 (0.818 to 0.870) | 35 | 0.722 (0.646 to 0.792) | 0.854 (0.769 to 0.950) |
| | | HCV | 260 | 0.737 (0.713 to 0.760) | 96 | 0.671 (0.625 to 0.715) | 0.910 (0.845 to 0.980) |
| Vargas et al., 2015[24] | Chile | HCV | 9 | 0.682 (0.118 to 0.996) | 2 | 0.536 (0.025 to 0.988) | 0.786 (0.209 to 2.954) |
| Vellopoulou et al., 2014[25] | Netherland | HCV | 23 | 0.730 (0.649 to 0.804) | 4 | 0.500 (0.302 to 0.698) | 0.685 (0.453 to 1.036) |
| Woo et al., 2012[26] | Canada | HBV | 79 | 0.880 (0.843 to 0.913) | 7 | 0.730 (0.379 to 0.962) | 0.830 (0.545 to 1.262) |
| Wright et al., 2006[27] | UK | HCV | 40 | 0.550 (0.444 to 0.654) | 64 | 0.450 (0.365 to 0.537) | 0.818 (0.624 to 1.073) |
| **Pooled (random-effects meta-analysis)** $\chi^2$=20.87; df = 10; p=0.022; $\tau^2$=0.004; $I^2$=52.1% | | | | | | | **0.879 (0.824 to 0.939)** |

*CLD = chronic liver disease; HBV = hepatitis B virus; HCV = hepatitis C virus*

#### 4.3. Hepatocellular carcinoma

We use evidence from Verma *et al.*[28] to account for the impact of early- and late-stage HCC diagnosis on HRQoL. They provide SF-36 measurements before and after diagnosis, distinguishing between people who developed stage I-II and stage III-IV tumours (according to the American Joint Committee on Cancer [AJCC] staging system – analogous to the T-stage of TNM). We map domain-level SF-36 data onto EQ-5D-3L values using model 6 from Ara and Brazier's algorithm[29].

Before HCC diagnosis, participants who went on to develop HCC had EQ-5D-3L utility of 0.545. Those who developed stage I/II HCC in the subsequent 2 years had EQ-5D-3L utility of 0.535 (equivalent to a utility multiplier of 0.982, compared with baseline); people with stage III/IV HCC had EQ-5D-3L utility of 0.480 (utility multiplier = 0.880, compared with baseline).

In the model, we apply these multipliers to the utility values for F2/F3/F4 fibrosis (which, in turn, we derive using a multiplier for fibrosis compared with expected utility in age- and sex-matched population; see main text). We assume the value for stage I/II HCC is relevant for people with BCLC 0/A disease, and the value for stage III/IV HCC applies to people with

BCLC B/C/D tumours. For people with underlying decompensated cirrhosis (i.e. BCLC stage D HCC), we also apply the multiplier described in *4.1*, above.

In probabilistic analyses, we account for uncertainty in these quantities by sampling values for the domain-level SF-36 scores and the associated SF-36–EQ-5D-3L mapping coefficients from independent normal distributions. Ideally, we would account for correlations in both steps in multivariate sampling; however, information on covariance is unavailable in either case.

#### eAppendix 5 State occupancy graphs for each starting state

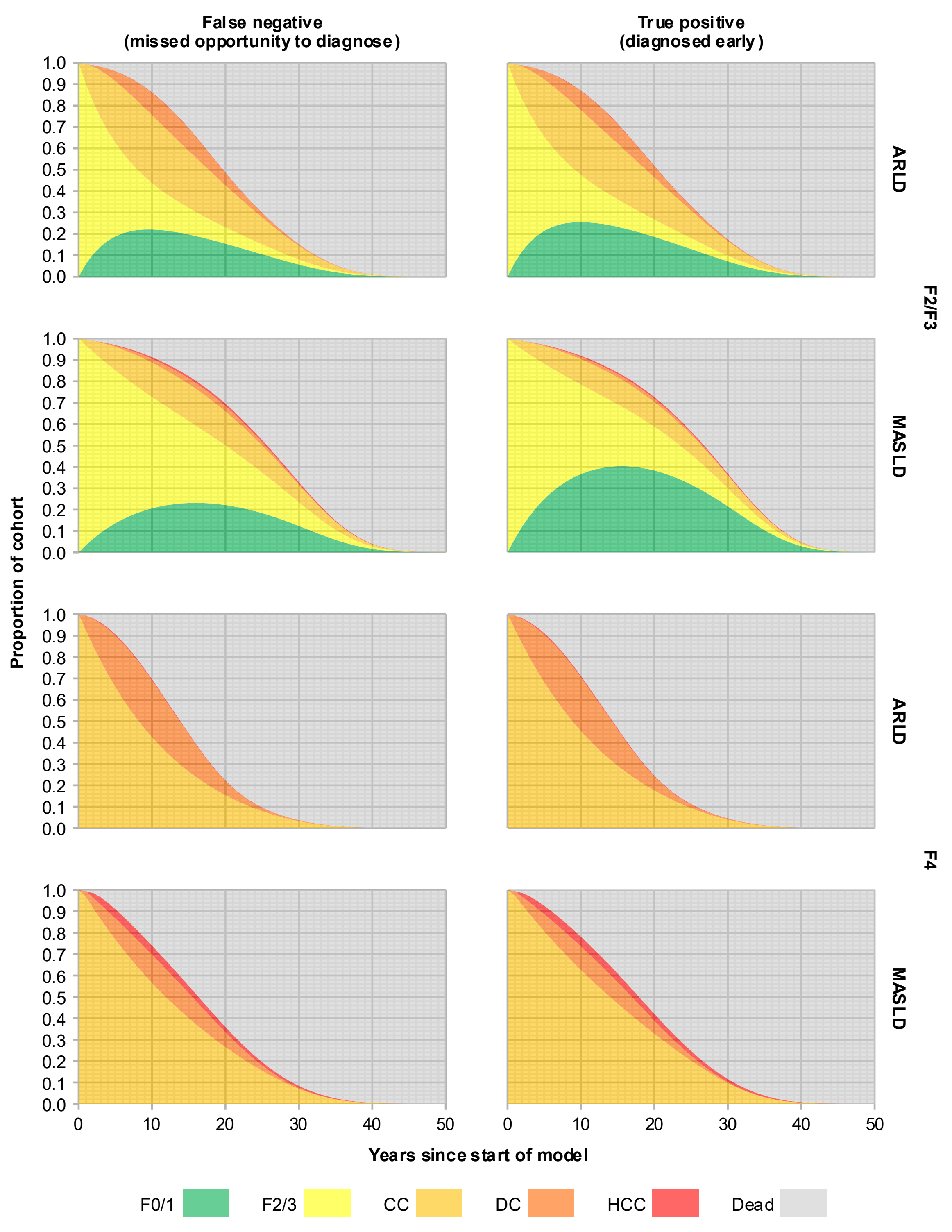

*ARLD = alcohol-related liver disease; CC = compensated cirrhosis; DC = decompensated cirrhosis; F0/1/2/3/4 = METAVIR fibrosis stage; HCC = hepatocellular carcinoma; MASLD = metabolic-dysfunction-associated steatotic liver disease*

*See Table 1 in main text for starting ages of each cohort.*

**Figure e2 State occupancy over time for people with significant fibrosis, according to aetiology (ARLD -v- MASLD) and diagnosis status (FN -v- TP)**

#### eAppendix 6 Cost effectiveness of hypothetical strategies for identifying people with liver disease

##### Table e10 Expected lifetime life-years, quality-adjusted life-years, and hepatology costs for people with different levels of fibrosis, according to whether their disease is detected

| | Absolute | | | Incremental, TP -v- FN | | | Maximum justifiable cost per TP identified[b] |
|---|---|---|---|---|---|---|---|
| | LYs | QALYs[a] | Costs[a] | LYs | QALYs[a] | Costs[a] | |
| **ARLD** | | | | | | | |
| Minimal fibrosis (F0/F1) | | | | | | | |
| TN or FP | 25.78 | 11.74 | £10,702 | | | | |
| Significant fibrosis (F2/F3) | | | | | | | |
| FN | 20.57 | 8.87 | £26,597 | | | | |
| TP | 21.21 | 9.06 | £27,490 | 0.64 | 0.20 | £892 | £3,009 |
| Compensated cirrhosis (F4) | | | | | | | |
| FN | 15.04 | 6.93 | £70,391 | | | | |
| TP | 15.52 | 7.10 | £71,708 | 0.47 | 0.17 | £1,316 | £2,182 |
| F2 or worse | | | | | | | |
| FN | 18.43 | 8.12 | £43,558 | | | | |
| TP | 19.01 | 8.30 | £44,614 | 0.58 | 0.19 | £1,056 | £2,689 |
| **MASLD** | | | | | | | |
| Minimal fibrosis (F0/F1) | | | | | | | |
| TN or FP | 29.17 | 13.01 | £1,702 | | | | |
| Significant fibrosis (F2/F3) | | | | | | | |
| FN | 25.20 | 10.12 | £6,123 | | | | |
| TP | 26.05 | 10.35 | £7,729 | 0.85 | 0.23 | £1,607 | £2,943 |
| Compensated cirrhosis (F4) | | | | | | | |
| FN | 17.34 | 7.55 | £31,860 | | | | |
| TP | 18.71 | 8.03 | £34,293 | 1.37 | 0.47 | £2,434 | £6,972 |
| F2 or worse | | | | | | | |
| FN | 23.70 | 9.63 | £11,033 | | | | |
| TP | 24.65 | 9.90 | £12,797 | 0.95 | 0.27 | £1,764 | £3,712 |
| **Weighted average** | | | | | | | |
| Minimal fibrosis (F0/F1) | | | | | | | |
| TN or FP | 28.47 | 12.75 | £3,551 | | | | |
| Significant fibrosis (F2/F3) | | | | | | | |
| FN | 23.60 | 9.69 | £13,179 | | | | |
| TP | 24.38 | 9.91 | £14,539 | 0.78 | 0.22 | £1,360 | £2,966 |
| Compensated cirrhosis (F4) | | | | | | | |
| FN | 15.99 | 7.19 | £54,556 | | | | |
| TP | 16.83 | 7.48 | £56,332 | 0.84 | 0.30 | £1,775 | £4,150 |
| F2 or worse | | | | | | | |
| FN | 21.38 | 8.96 | £25,261 | | | | |
| TP | 22.18 | 9.20 | £26,743 | 0.80 | 0.24 | £1,482 | £3,312 |

[a] *Discounted 3.5% per year*

[b] *When QALYs are valued at £20,000 each. Any case-identification strategy that finds people with the specified level of fibrosis at a cost per true-positive less than this will have positive net benefit (i.e. an ICER better than £20,000/QALY, compared with no case-identification).*

*ARLD = alcohol-related liver disease; FN = false negative; FP = false positive; LYs = life-years; MASLD = metabolic dysfunction-associated liver disease; QALYs = quality-adjusted life-years; TN = true negative; TP = true positive*

*Table e10* shows expected lifetime costs and QALY for true-positives and false-negatives in ARLD, MASLD, and mixed populations. There is no population in which detection of disease leads to net cost-savings. This is because, although we reduce downstream healthcare costs (decompensation; HCC) by slowing progression of liver-disease, expenditure on lifestyle intervention and ongoing follow-up slightly outweigh this saving. However, there is substantial health-gain associated with detection (approximately 0.2 QALYs for people with ARLD; approximately 0.3 QALYs for people with MASLD). If we value QALYs at £20,000 each (as per NICE's lower cost-effectiveness threshold), any programme that detects significant liver-disease at a cost of less than £3,300 per case would generate positive net benefit.

Armed with this information, we can estimate the cost-effectiveness of any strategy for which we know sensitivity, specificity and up-front costs (see *Figure e3*).

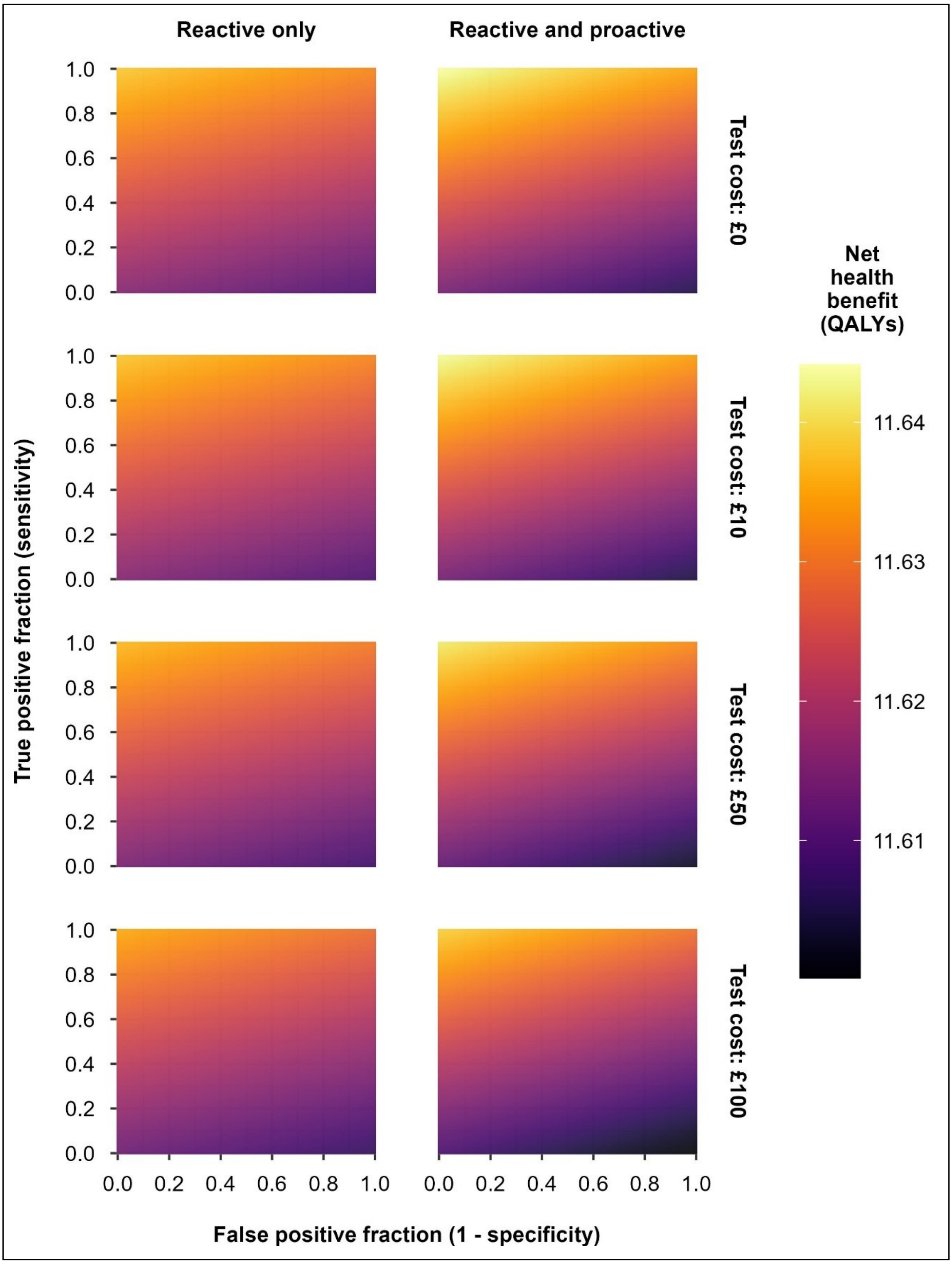

**Figure e3** **Value for money for risk-stratification strategies with any combination of sensitivity and specificity, at a range of possible test costs**

#### eAppendix 7 Probabilistic sensitivity analysis

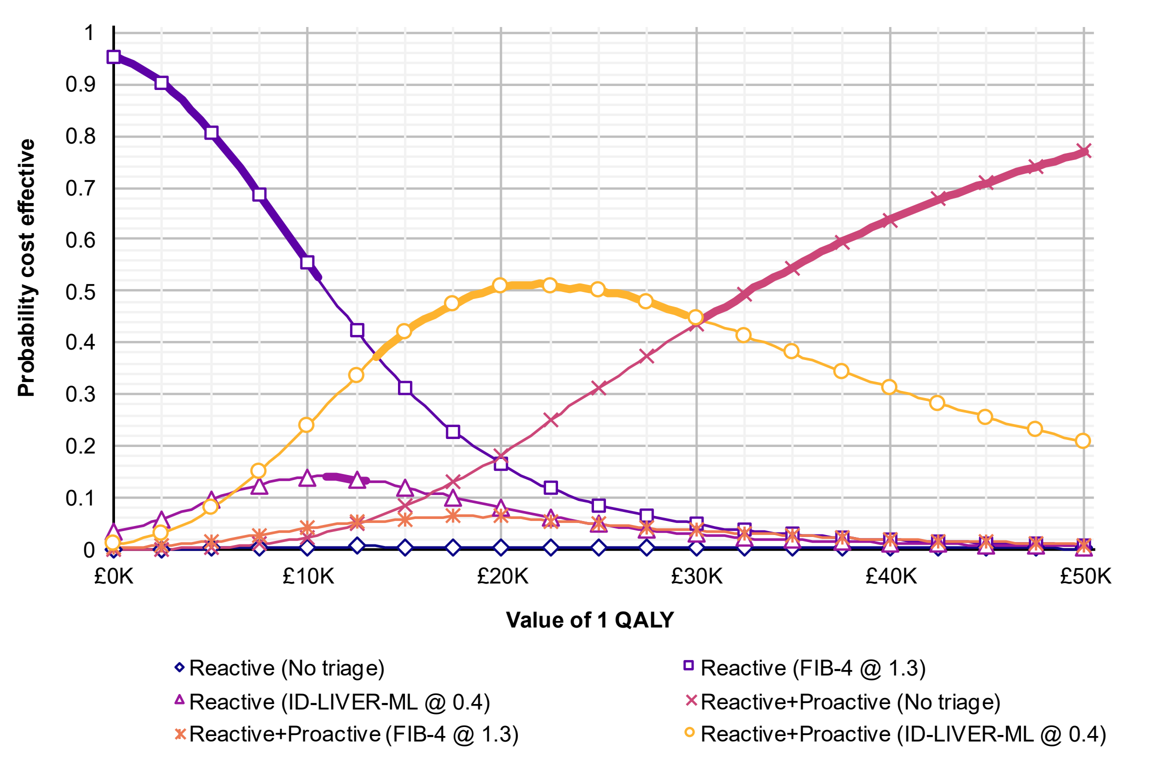

*Bold line shows cost-effectiveness acceptability frontier*

**Figure e4 Cost-effectiveness acceptability curve and frontier**

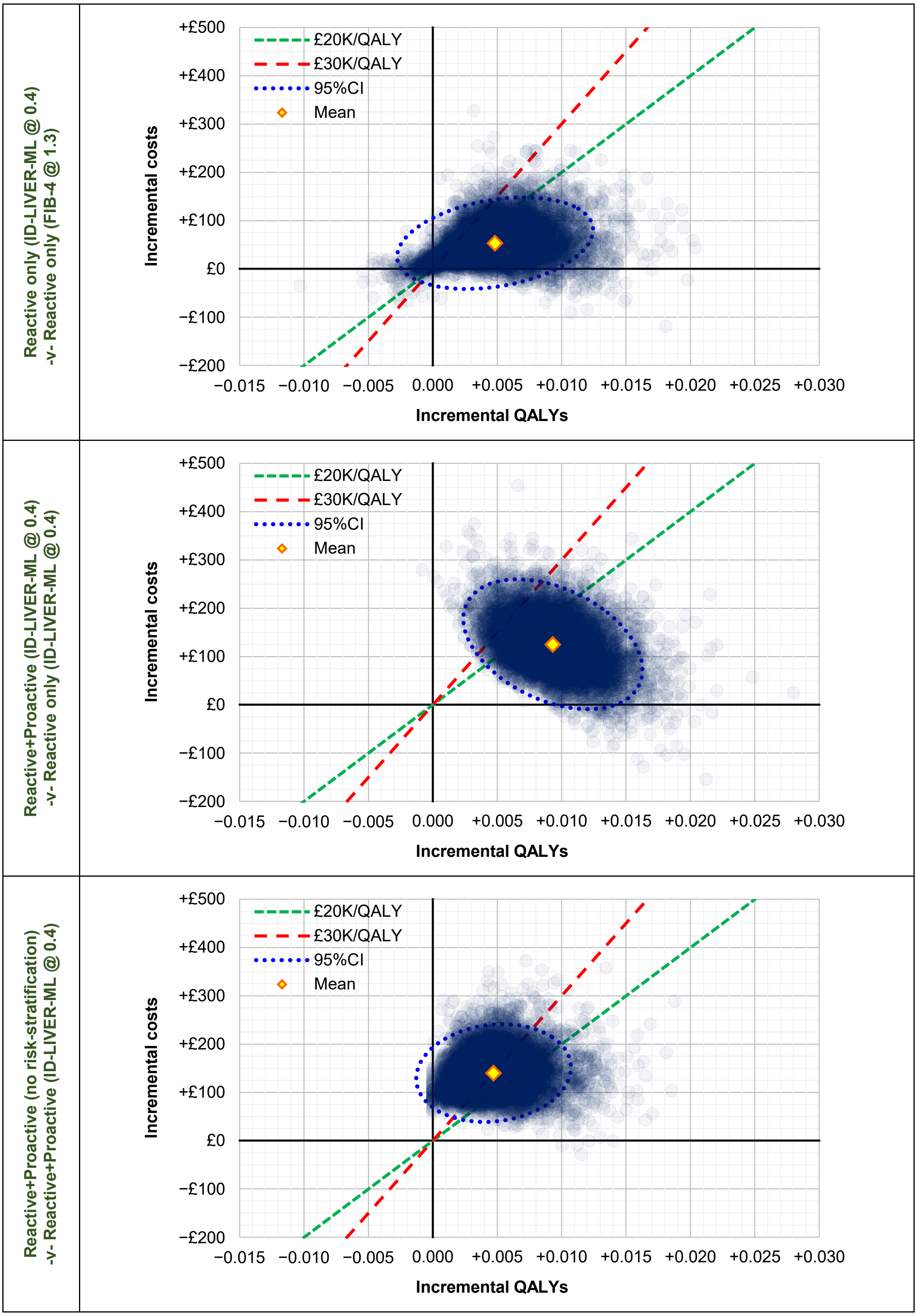

**Figure e5** **Scatterplots showing incremental costs and QALYs for key pairwise comparisons in probabilistic analysis (10,000 iterations)**

#### eAppendix 8 One-way sensitivity analysis

We focus, here, on the three sequential pairwise comparisons on the cost-effectiveness frontier in our base case, as these are the comparisons on which our decision uncertainty rests:

1. Compared with the cheapest thing we can do (FIB-4 @ 1.3 in the reactive-only population), is it worth paying extra money to gain more QALYs by using ID-LIVER-ML @ 0.4 instead (*Figure e6*)?
2. Is it good value to extend coverage from the reactive-only population to introduce proactive case-finding in the community (*Figure e7*)?
3. Can we afford to remove risk-stratification so that secondary-care services review all people identified as at risk of CLD, which is guaranteed to maximise QALYs, but will also incur greater costs (*Figure e8*)?

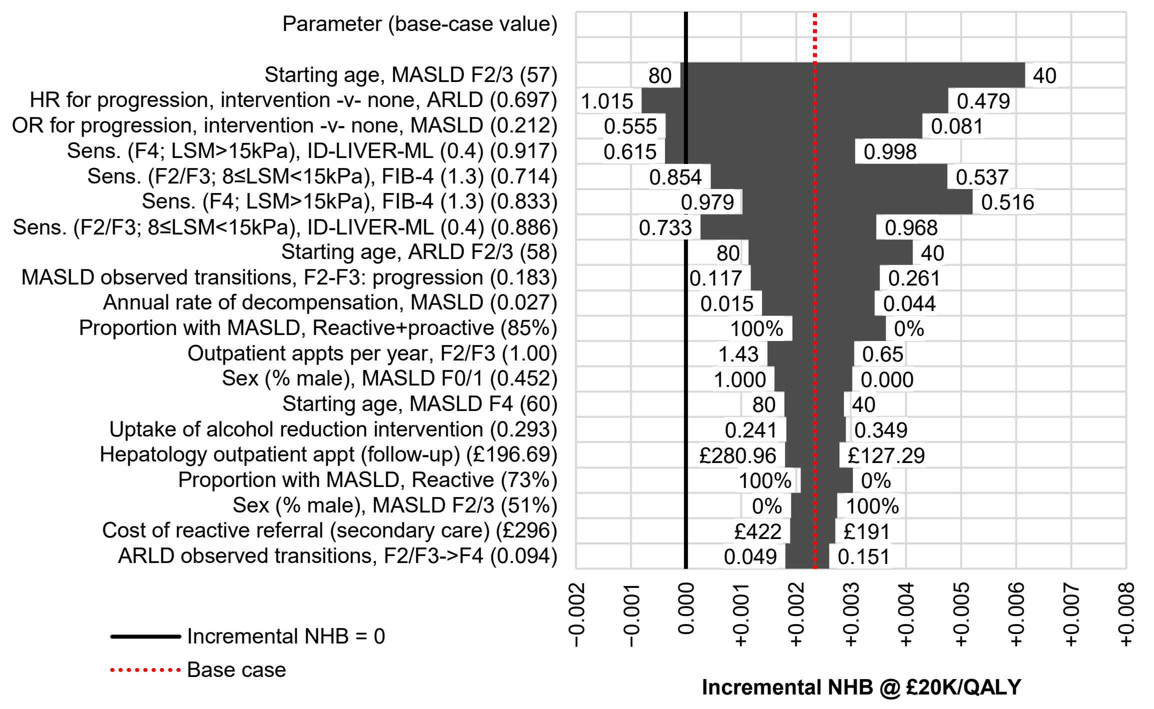

*Appt. = appointment; ARLD = alcohol-related liver disease; F0/1/2/3/4 = METAVIR fibrosis stage; HR = hazard ratio; LSM = liver stiffness measurement; MASLD = metabolic-dysfunction-associated steatotic liver disease; OR = odds ratio; Sens. = sensitivity; Spec. = specificity*

*Positive incremental NHB values favour ID-LIVER-ML; negative values favour FIB-4. Most influential 20 parameters shown.*

**Figure e6 One-way sensitivity analysis, reactive-only (ID-LIVER-ML @ 0.4) -v- reactive-only (FIB-4 @ 1.3)**

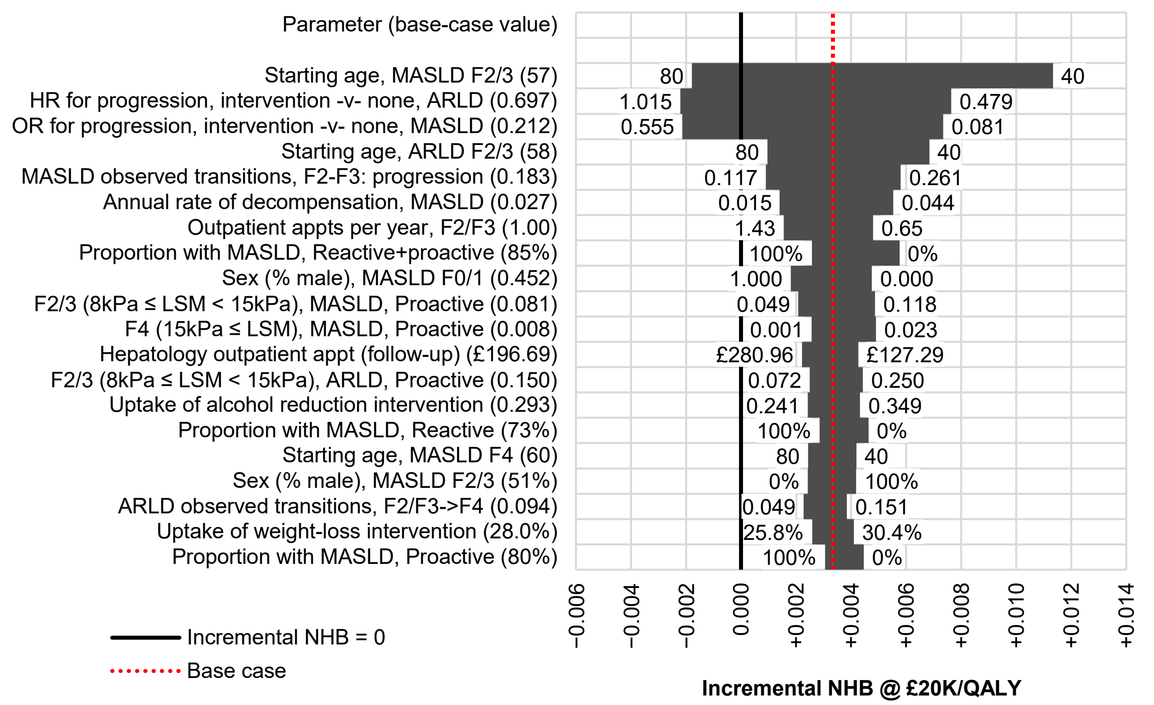

*Appt. = appointment; ARLD = alcohol-related liver disease; F0/1/2/3/4 = METAVIR fibrosis stage; HR = hazard ratio; LSM = liver stiffness measurement; MASLD = metabolic-dysfunction-associated steatotic liver disease; OR = odds ratio; Sens. = sensitivity; Spec. = specificity*

*Positive incremental NHB values favour ID-LIVER-ML; negative values favour FIB-4. Most influential 20 parameters shown.*

**Figure e7 One-way sensitivity analysis, reactive+proactive (ID-LIVER-ML @ 0.4) -v- reactive-only (ID-LIVER-ML @ 0.4)**

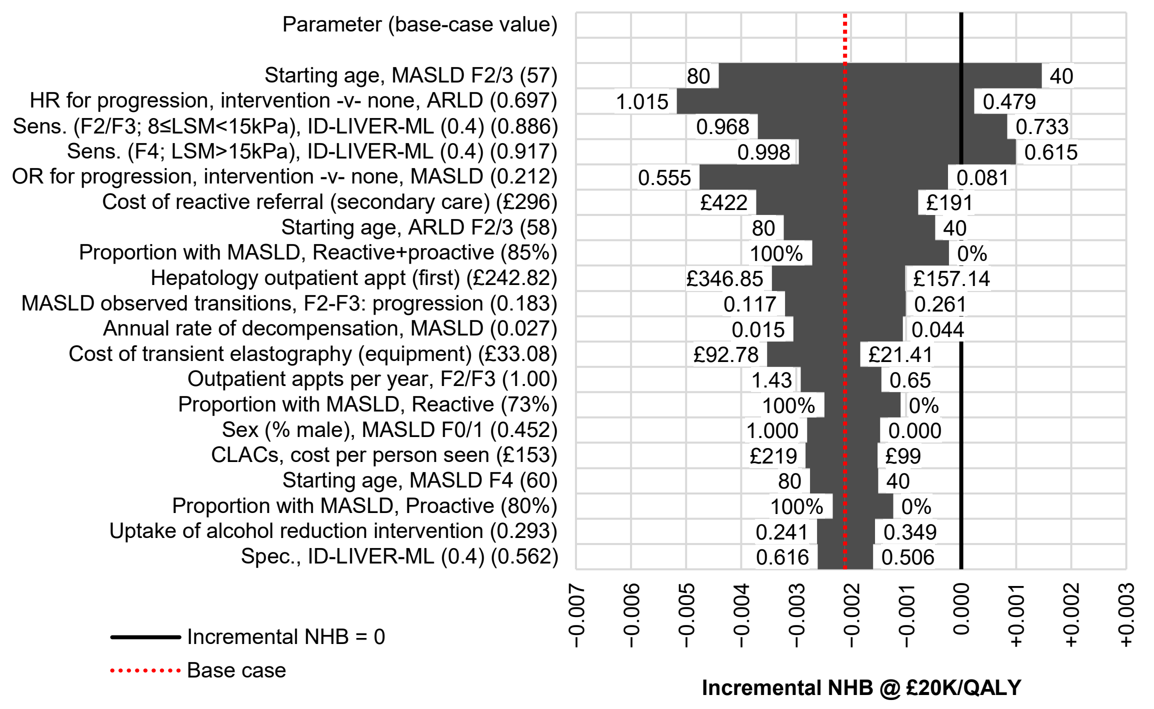

*Appt. = appointment; ARLD = alcohol-related liver disease; F0/1/2/3/4 = METAVIR fibrosis stage; HR = hazard ratio; LSM = liver stiffness measurement; MASLD = metabolic-dysfunction-associated steatotic liver disease; OR = odds ratio; Sens. = sensitivity; Spec. = specificity*

*Positive incremental NHB values favour ID-LIVER-ML; negative values favour FIB-4. Most influential 20 parameters shown.*

**Figure e8 One-way sensitivity analysis, reactive+proactive (no risk-stratification) -v- reactive+proactive (ID-LIVER-ML @ 0.4)**

### eAppendix 9 Scenario analyses

#### 9.1. Removing risk-stratification from proactive case-finding

When we simulate a combination of proactive case-finding and risk-stratification (with FIB-4 or ID-LIVER-ML) in our base case, we effectively assume that data will be available to calculate risk scores for the people identified as at risk of CLD in digital searches of primary care records. This may not always be true, as both risk-stratification tools require reasonably up-to-date inputs (e.g. liver function tests). In the reactive setting, it is reasonable to assume these will always be available, as the referral comes following consultation with primary care clinicians who will invariably order any necessary tests and provide relevant information. However, people in the proactive population, whose risk factors have been identified without any particular interaction with healthcare providers, may not have up-to-date information from which to calculate a risk score. Therefore, we undertook a scenario analysis in which we assumed risk-stratification tools are only relevant to the reactive population, while people in the proactive population will always have to attend a CLAC in order to undergo assessment. Figure e9 illustrates our revised approach to defining the available strategies (cf. *Figure 1* in the main text).

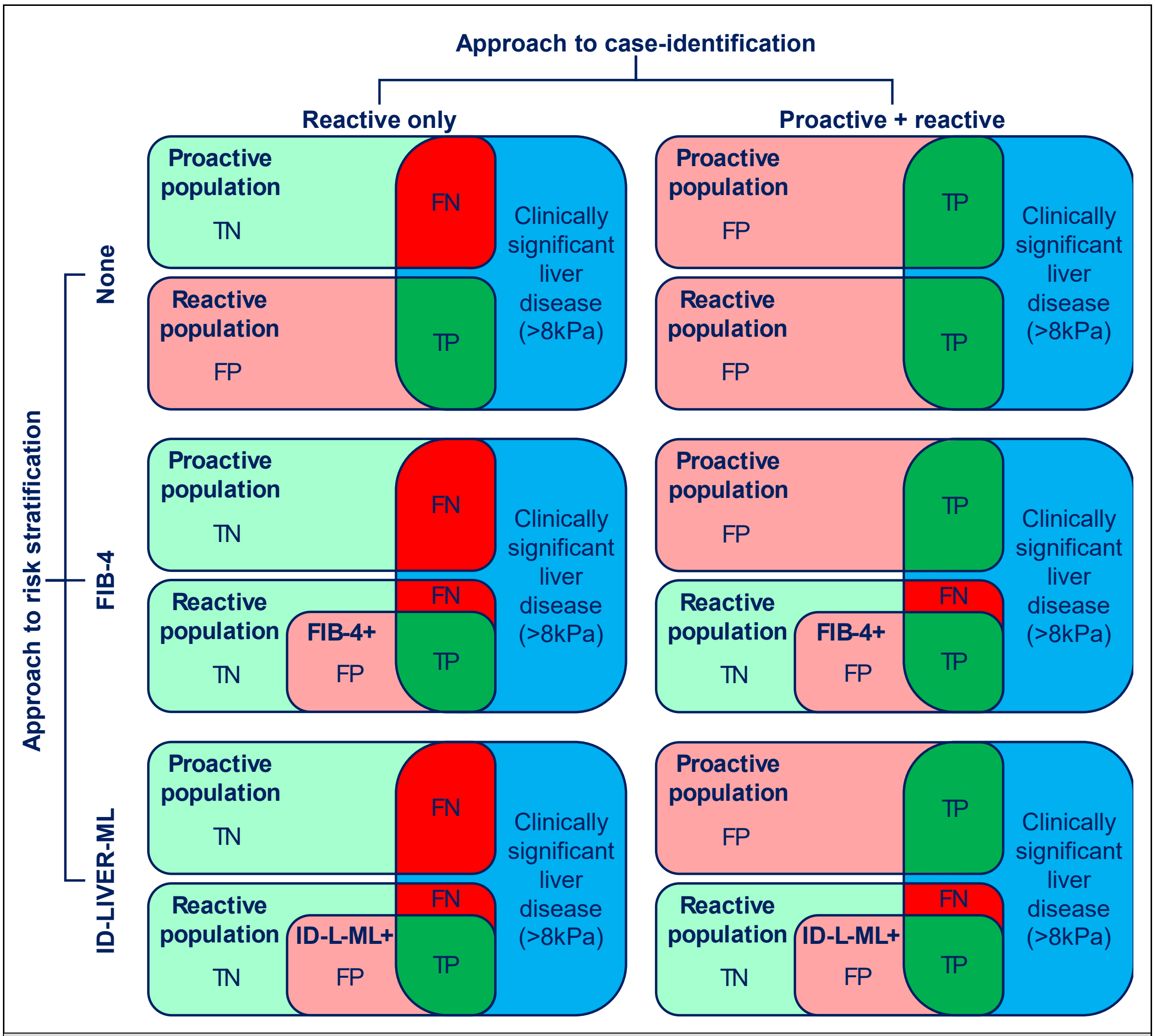

*FN = false negative; FP = false negative; ID-L-ML = ID-LIVER machine-learning risk-prediction model; TN = true negative; TP = true positive*

**Figure e9 Illustration of the six strategies evaluated, combining two approaches to case-identification and three approaches to risk-stratification, when risk-stratification is not possible for the proactive population**

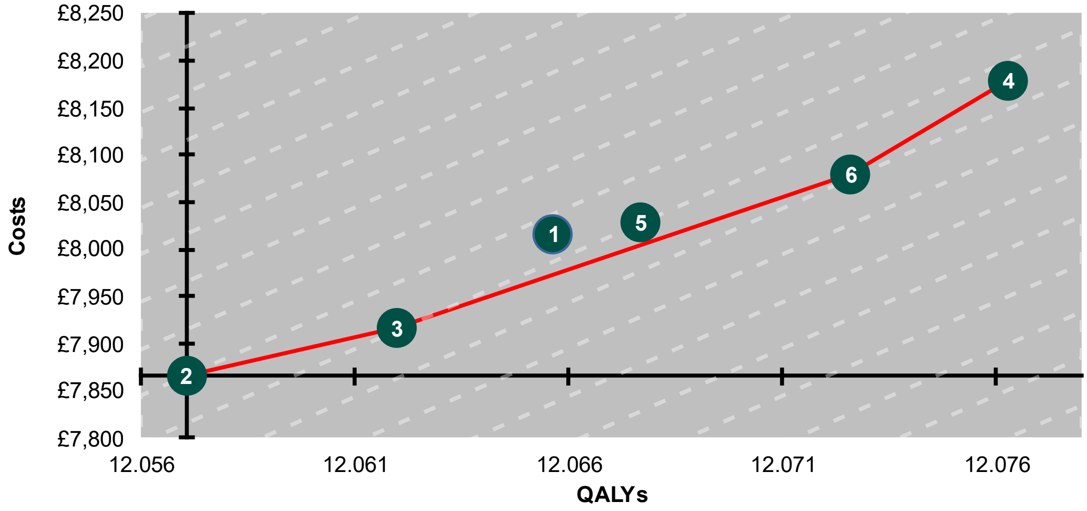

(1) Reactive (No triage)
(2) Reactive (FIB-4 @ 1.3)
(3) Reactive (ID-LIVER-ML @ 0.4)
(4) Reactive+Proactive (No triage)
(5) Reactive+Proactive (FIB-4 @ 1.3)
(6) Reactive+Proactive (ID-LIVER-ML @ 0.4)

| Strategy | | Absolute | | Incremental | | | Net health benefit | |
|---|---|---|---|---|---|---|---|---|
| Case-finding | Risk-stratification (threshold) | Costs (£) | Effects (QALYs) | Costs (£) | Effects (QALYs) | ICER (£/QALY) | £20K/ QALY | £30K/ QALY |
| Reactive | FIB-4 (1.3) | £7,865 | 12.057 | | | | 11.664 | 11.795 |
| Reactive | ID-LIVER-ML (0.4) | £7,917 | 12.062 | £52 | 0.005 | £10,498 | 11.666 | 11.798 |
| Reactive | None | £8,015 | 12.066 | £98 | 0.004 | ext. dom. | 11.665 | 11.798 |
| Reactive+Proactive | FIB-4 (1.3) | £8,029 | 12.068 | £111 | 0.006 | ext. dom. | 11.666 | 11.800 |
| Reactive+Proactive | ID-LIVER-ML (0.4) | £8,080 | 12.073 | £163 | 0.011 | £15,372 | 11.669 | 11.803 |
| Reactive+Proactive | None | £8,179 | 12.076 | £98 | 0.004 | £26,729 | 11.667 | 11.804 |

*Dashed line in background shows iso-net-benefit gradient equal to £20,000 per QALY. If the imaginary line between any two points is shallower than this gradient, the option on the right (generating most QALYs) represents good value; if it is steeper, the option on the left is better value, assuming we value QALYs at £20,000 each.*

**Figure e10 Scenario analysis removing risk-stratification from proactive case-finding: cost-effectiveness results**

#### 9.2. Limiting effectiveness of lifestyle interventions to single transitions that most closely reflect context in which the underlying studies collected data

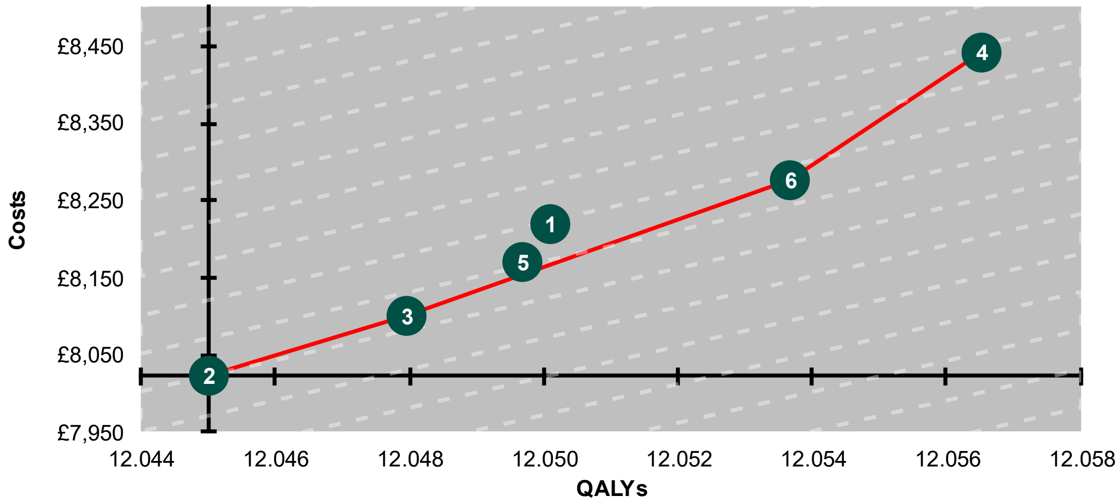

(1) Reactive (No triage)
(2) Reactive (FIB-4 @ 1.3)
(3) Reactive (ID-LIVER-ML @ 0.4)
(4) Reactive+Proactive (No triage)
(5) Reactive+Proactive (FIB-4 @ 1.3)
(6) Reactive+Proactive (ID-LIVER-ML @ 0.4)

| Strategy | | Absolute | | Incremental | | | Net health benefit | |
|---|---|---|---|---|---|---|---|---|
| Case-finding | Risk-stratification (threshold) | Costs (£) | Effects (QALYs) | Costs (£) | Effects (QALYs) | ICER (£/QALY) | £20K/ QALY | £30K/ QALY |
| Reactive | FIB-4 (1.3) | £8,022 | 12.045 | | | | 11.644 | 11.778 |
| Reactive | ID-LIVER-ML (0.4) | £8,101 | 12.048 | £79 | 0.003 | £26,893 | 11.643 | 11.778 |
| Reactive+Proactive | FIB-4 (1.3) | £8,170 | 12.050 | £69 | 0.002 | ext. dom. | 11.641 | 11.777 |
| Reactive | None | £8,220 | 12.050 | £119 | 0.002 | ext. dom. | 11.639 | 11.776 |
| Reactive+Proactive | ID-LIVER-ML (0.4) | £8,277 | 12.054 | £175 | 0.006 | £30,713 | 11.640 | 11.778 |
| Reactive+Proactive | None | £8,443 | 12.057 | £166 | 0.003 | £58,270 | 11.634 | 11.775 |

*Dashed line in background shows iso-net-benefit gradient equal to £20,000 per QALY. If the imaginary line between any two points is shallower than this gradient, the option on the right (generating most QALYs) represents good value; if it is steeper, the option on the left is better value, assuming we value QALYs at £20,000 each.*

**Figure e11 Scenario analysis limiting effectiveness of lifestyle interventions to single transitions that most closely reflect context in which the underlying studies collected data: cost-effectiveness results**

#### 9.3. Assuming health-related quality of life changes with F-stage in people with predominantly asymptomatic disease

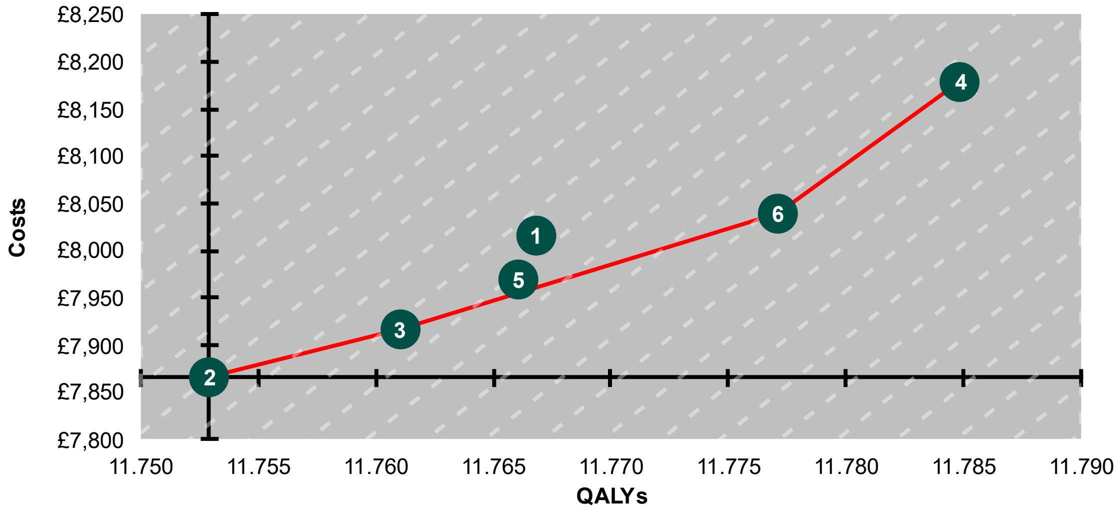

(1) Reactive (No triage)
(2) Reactive (FIB-4 @ 1.3)
(3) Reactive (ID-LIVER-ML @ 0.4)
(4) Reactive+Proactive (No triage)
(5) Reactive+Proactive (FIB-4 @ 1.3)
(6) Reactive+Proactive (ID-LIVER-ML @ 0.4)

| Strategy | | Absolute | | Incremental | | | Net health benefit | |
|---|---|---|---|---|---|---|---|---|
| Case-finding | Risk-stratification (threshold) | Costs (£) | Effects (QALYs) | Costs (£) | Effects (QALYs) | ICER (£/QALY) | £20K/ QALY | £30K/ QALY |
| Reactive | FIB-4 (1.3) | £7,865 | 11.753 | | | | 11.360 | 11.491 |
| Reactive | ID-LIVER-ML (0.4) | £7,917 | 11.761 | £52 | 0.008 | £6,389 | 11.365 | 11.497 |
| Reactive+Proactive | FIB-4 (1.3) | £7,970 | 11.766 | £52 | 0.005 | ext. dom. | 11.368 | 11.500 |
| Reactive | None | £8,015 | 11.767 | £98 | 0.006 | ext. dom. | 11.366 | 11.500 |
| Reactive+Proactive | ID-LIVER-ML (0.4) | £8,040 | 11.777 | £123 | 0.016 | £7,624 | 11.375 | 11.509 |
| Reactive+Proactive | None | £8,179 | 11.785 | £139 | 0.008 | £17,810 | 11.376 | 11.512 |

*Dashed line in background shows iso-net-benefit gradient equal to £20,000 per QALY. If the imaginary line between any two points is shallower than this gradient, the option on the right (generating most QALYs) represents good value; if it is steeper, the option on the left is better value, assuming we value QALYs at £20,000 each.*

**Figure e12 Scenario analysis assuming health-related quality of life changes with F-stage in people with predominantly asymptomatic disease: cost-effectiveness results**

#### 9.4. Costing proactive case-finding using costs assigned during ID-LIVER research project

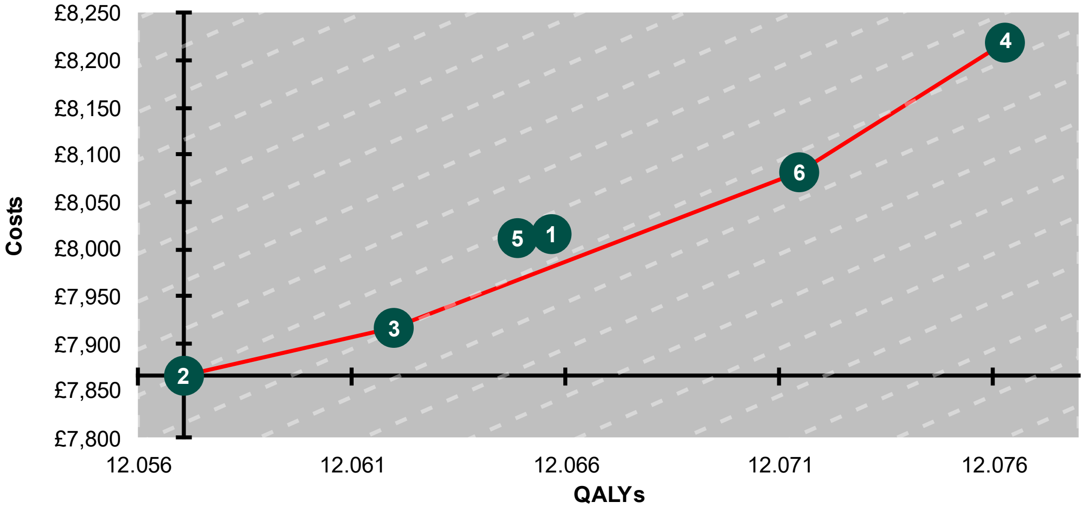

(1) Reactive (No triage)
(2) Reactive (FIB-4 @ 1.3)
(3) Reactive (ID-LIVER-ML @ 0.4)
(4) Reactive+Proactive (No triage)
(5) Reactive+Proactive (FIB-4 @ 1.3)
(6) Reactive+Proactive (ID-LIVER-ML @ 0.4)

| Strategy | | Absolute | | Incremental | | | Net health benefit | |
|---|---|---|---|---|---|---|---|---|
| **Case-finding** | **Risk-stratification (threshold)** | **Costs (£)** | **Effects (QALYs)** | **Costs (£)** | **Effects (QALYs)** | **ICER (£/QALY)** | **£20K/ QALY** | **£30K/ QALY** |
| Reactive | FIB-4 (1.3) | £7,865 | 12.057 | | | | 11.664 | 11.795 |
| Reactive | ID-LIVER-ML (0.4) | £7,917 | 12.062 | £52 | 0.005 | £10,498 | 11.666 | 11.798 |
| Reactive+Proactive | FIB-4 (1.3) | £8,011 | 12.065 | £94 | 0.003 | ext. dom. | 11.664 | 11.798 |
| Reactive | None | £8,015 | 12.066 | £98 | 0.004 | ext. dom. | 11.665 | 11.798 |
| Reactive+Proactive | ID-LIVER-ML (0.4) | £8,081 | 12.071 | £164 | 0.009 | £17,340 | 11.667 | 11.802 |
| Reactive+Proactive | None | £8,220 | 12.076 | £139 | 0.005 | £28,753 | 11.665 | 11.802 |

*Dashed line in background shows iso-net-benefit gradient equal to £20,000 per QALY. If the imaginary line between any two points is shallower than this gradient, the option on the right (generating most QALYs) represents good value; if it is steeper, the option on the left is better value, assuming we value QALYs at £20,000 each.*

**Figure e13 Scenario analysis using costs assigned to proactive case-finding during ID-LIVER research project**

#### eAppendix 10 Threshold analyses

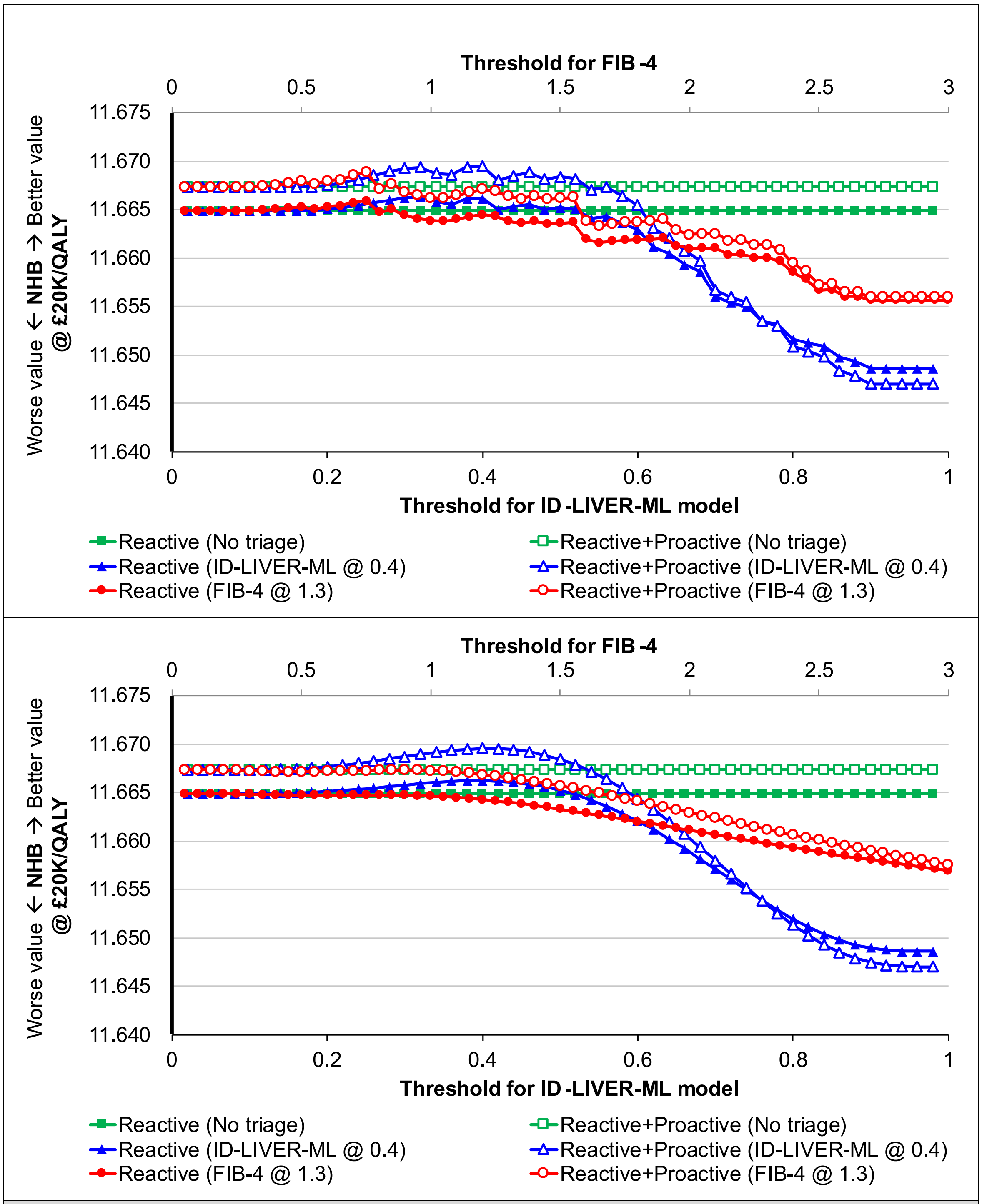

*NHB @ £20K/QALY = net health benefit when we value quality-adjusted life-years at £20,000 each*

*Upper panel shows results when we use patient-level data to estimate diagnostic accuracy; lower panel shows results using a beta regression fitted to those data*

**Figure e14 Threshold analysis: relationship between diagnostic threshold (cutoff) for FIB-4 and ID-LIVER-ML model and value for money (net health benefit)**

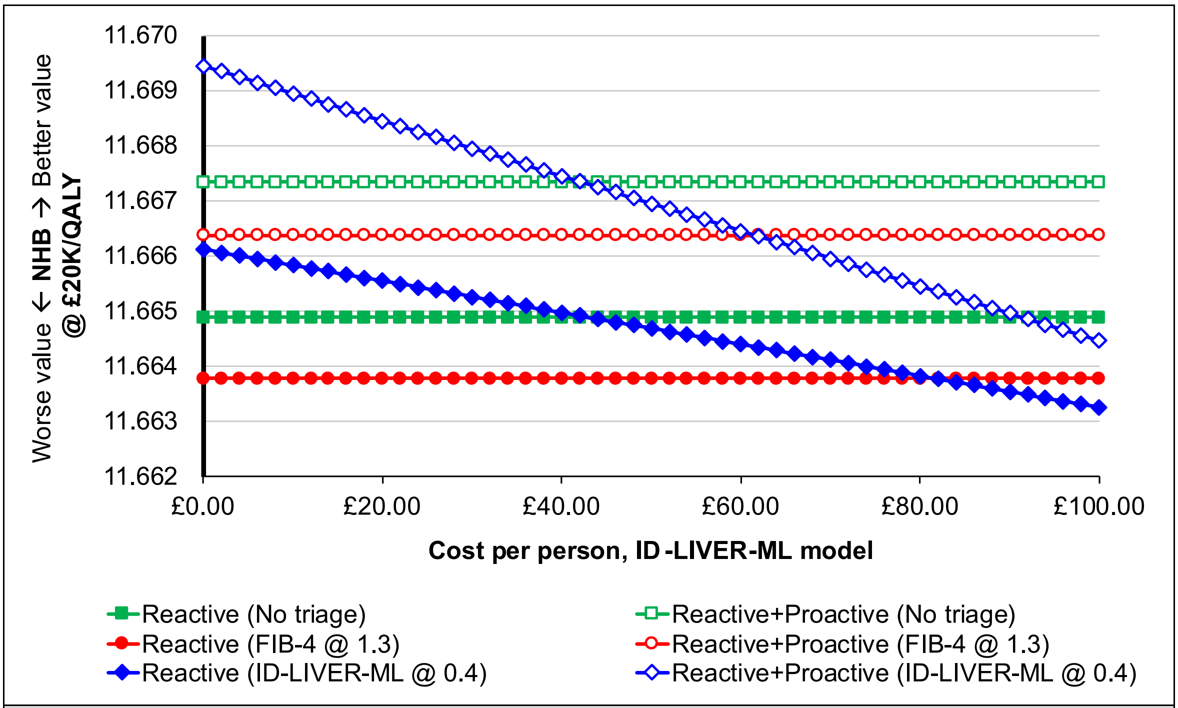

*NHB @ £20K/QALY = net health benefit when we value quality-adjusted life-years at £20,000 each*

**Figure e15 Threshold analysis: relationship between cost per person of ID-LIVER-ML model and value for money (net health benefit)**

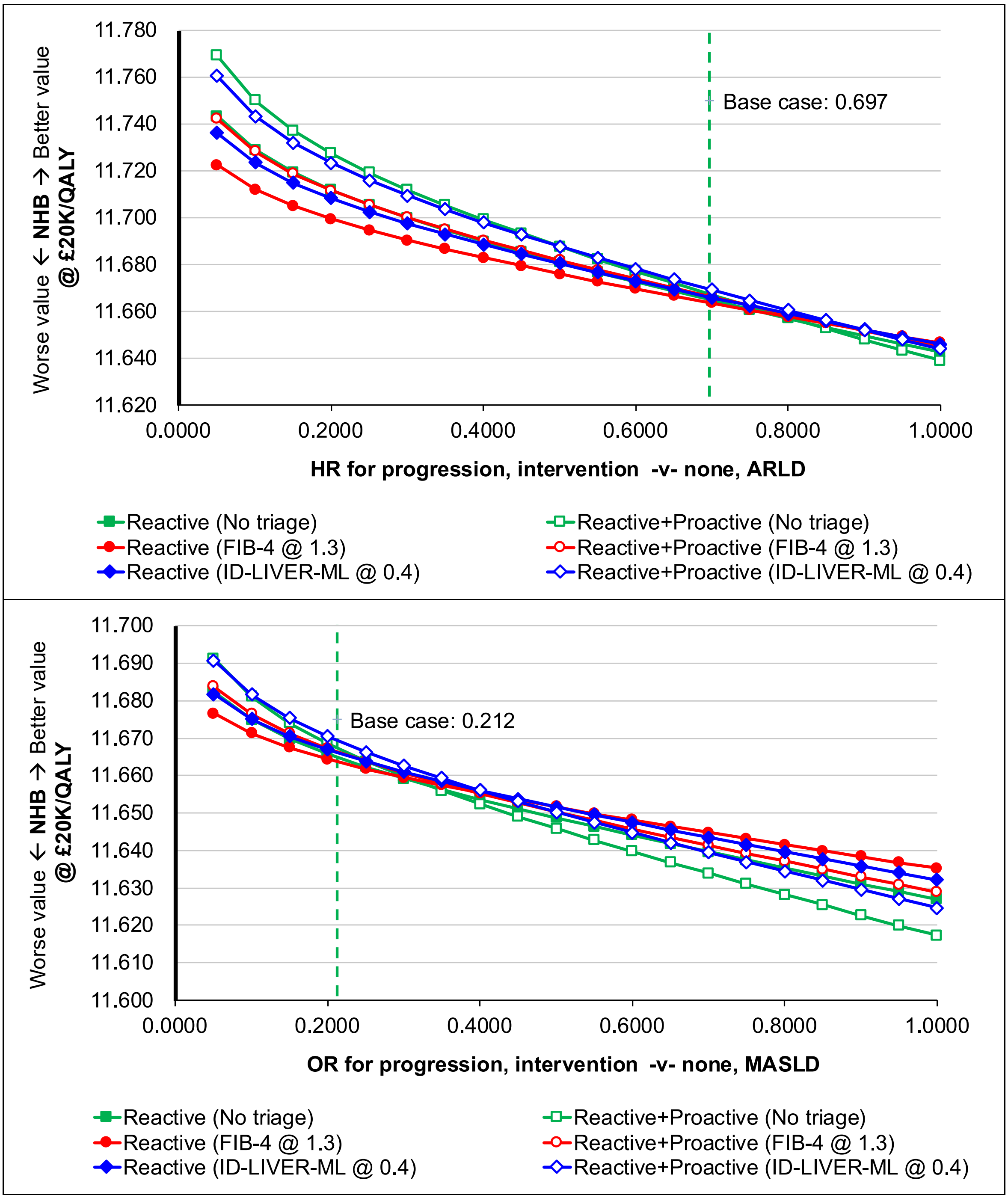

*ARLD = alcohol-related liver disease; MASLD = metabolic-dysfunction-associated steatotic liver disease;*
*NHB @ £20K/QALY = net health benefit when we value quality-adjusted life-years at £20,000 each*

*Upper panel shows results when varying effectiveness of lifestyle intervention for people with ARLD.*

- *Reactive+Proactive (No triage) generates greatest net benefit when HR ≤ 0.45*
- *Reactive+Proactive (ID-LIVER-ML @ 0.4) generates greatest net benefit when 0.5 ≤ HR ≤ 0.85*
- *Reactive (ID-LIVER-ML @ 0.4) generates greatest net benefit when HR = 0.90*
- *Reactive (FIB-4 @ 1.3) generates greatest net benefit when HR ≥ 0.95*

*Lower panel shows results when varying effectiveness of lifestyle intervention for people with MASLD.*

- *Reactive+Proactive (No triage) generates greatest net benefit when HR = 0.05*
- *Reactive+Proactive (ID-LIVER-ML @ 0.4) generates greatest net benefit when 0.05 < HR ≤ 0.4*
- *Reactive (ID-LIVER-ML @ 0.4) generates greatest net benefit when HR = 0.45*
- *Reactive (FIB-4 @ 1.3) generates greatest net benefit when HR ≥ 0.50*

**Figure e16 Threshold analysis: relationship between effectiveness of lifestyle interventions and value for money (net health benefit)**
